# AutoReporter: Development of an artificial intelligence tool for automated assessment of research reporting guideline adherence

**DOI:** 10.1101/2025.04.18.25326076

**Authors:** David Chen, Patrick Li, Ealia Khoshkish, Seungmin Lee, Tony Ning, Umair Tahir, Henry CY Wong, Michael SF Lee, Srinivas Raman

## Abstract

**Objective:** To develop AutoReporter, a large-language-model system that automates evaluation of adherence to research reporting guidelines.

**Materials and Methods:** Eight prompt-engineering and retrieval strategies coupled with reasoning and general-purpose LLMs were benchmarked on the SPIRIT-CONSORT-TM corpus. The top-performing approach, AutoReporter, was validated on BenchReport, a novel benchmark dataset of expert-rated reporting guideline assessments from 10 systematic reviews.

**Results:** AutoReporter, a zerolZlshot, nolZlretrieval prompt coupled with the o3-mini reasoning LLM, demonstrated optimal accuracy (CONSORT: 90.09%; SPIRIT: 92.07%), run-time (CONSORT: 617.26 seconds; SPIRIT: 544.51 seconds), and cost (CONSORT: 0.68 USD; SPIRIT: 0.65 USD). AutoReporter achieved a mean accuracy of 91.8% and substantial agreement (Cohen’s κ>0.6) with expert ratings from the BenchReport benchmark.

**Discussion:** Structured prompting alone can match or exceed fine-tuned domain models while forgoing manually annotated corpora and computationally intensive training.

**Conclusion:** LLMs can feasibly automate reporting guideline adherence assessments for scalable quality control in scientific research reporting. AutoReporter is publicly accessible at https://autoreporter.streamlit.app.

## Background and Significance

Concerns about scientific reproducibility have underscored the importance of comprehensive research reporting ^1,2^. Consensus guidelines such as CONSORT for randomized trials ^3^ and SPIRIT for trial protocols ^4^ aimed to standardize methodological reporting, yet recent audits report modest post-guideline improvements and persistent omissions across clinical research guidelines ^5,6^ and emerging artificial intelligence applications ^7^. There remains a translational research gap between theoretical endorsement and widespread adoption of reporting guidelines.

A significant barrier is the labor-intensive and subjective nature of assessing guideline adherence within a backdrop of rapidly expanding volumes of biomedical literature and novel extensions and versions of established reporting guidelines ^8,9^. To address this issue, expert-led editorial intervention can improve reporting completeness but is difficult to scale ^10^. Rule- and keyword-driven natural language processing (NLP) methods can detect CONSORT items with above 90% accuracy ^11^, but are limited in contextual and semantic understanding necessary to generalize well across diverse research texts and guidelines. Machine learning (ML) classifiers show promise in classifying scientific article types ^12^ and assessing risk of bias in systematic review contexts ^13^, motivating the application of modern ML-derived large language models (LLM) with near-human NLP competencies to automate reporting guideline assessment ^14,15^.

Domain-specific LLMs such as BioBERT^16^ and PubMedBERT ^17^ have been tested in pilot case studies of guideline assessment but require large-scale resources and extensive, annotated training data. Zero-shot applications of general-purpose LLMs, such as the popular Generative Pre-Trained Transformer (GPT), have yielded promising results, yet retrievalCaugmented prompt methods and reasoning-based LLMs remain largely untested across multiple guidelines and study designs ^18–20^. We developed AutoReporter, an LLM system that automates reporting guideline adherence of full-text manuscripts, delivering realCtime reporting feedback for authors and scalable bulk screening of reporting adherence for journal editorial teams.

## Materials and Methods

### Datasets

The SPIRIT-CONSORT-TM corpus contains 100 English RCT reports and 100 English RCT protocols, each annotated with reporting adherence for 83 SPIRIT and CONSORT items ^21^ (Supplementary Figure 1). The SPIRIT-CONSORT-TM corpus ^21^ operationalizes reporting guidelines at the level of 83 granular elements. These correspond to the 25 top-level CONSORT checklist items and the 33 SPIRIT checklist items, expanded into sub-elements for annotation. Out of the 83 items from both guidelines, four items were applicable to SPIRIT only, eleven items were applicable to CONSORT only, and 78 items were applicable to both guidelines. BenchReport is a novel benchmark dataset that aggregates 10,710 human ratings for 506 English articles included in 10 systematic reviews, spanning 10 reporting guidelines, 7 clinical domains, and 6 article types ^22–29^ (Supplementary Table 1, Supplementary Figure 1). The case study evaluation dataset comprises 10 systematic reviews first-authored or supervised by H.C.Y.W. (n=5) or M.S.F.L (n=5). Dataset methods are described in the Datasets section of Supplement.

### LLM and Prompt Method Design

We evaluated two general-purpose (gpt-4o, gpt-4o-mini) ^30^ and two reasoning (o1, o3-mini) ^31^ LLMs via the OpenAI API with default parameters. Inputs comprised retrieved article text, guideline item number, item description, and with fewCshot prompts, examples of reported items. ZeroCshot and fewCshot prompts that included one to three examples from the SPIRITCCONSORTCTM annotation guide were tested with four retrievalCaugmented generation (RAG) modes: no RAG, BM25, Contextual (Supplementary Figure 2), and Hybrid RAG methods. The input prompt and expected output from AutoReporter is shown in Supplementary Table 2. The output of AutoReporter included the binary rating of one item’s reporting status (0: not wholly reported; 1: wholly reported) with quoted evidence, and if absent, a suggestion to address the reporting deficiency. LLM methods described in LLM Design and Prompt Method Design sections of Supplement.

### Evaluation Procedure

The template prompt structure, including wording and text ordering, was empirically tested on the CONSORT Train split (n = 10). Eight prompt and RAG method combinations were validated on CONSORT Validate (n = 20) using the general-purpose gptC4oCmini and reasoning gptCo3Cmini LLMs. The top-performing zero-shot, no RAG retrieval prompt was tested on CONSORT Test (n = 20) and SPIRIT Test (n = 20) with the general-purpose gpt-4o and gptC4oCmini as well as reasoning o1 and o3Cmini LLMs to compare with the state of the art PubMedBERT method. The top-performing zero-shot, no RAG retrieval prompt with the reasoning o3-mini LLM identified from the CONSORT Validate test, known as AutoReporter, was tested on the BenchReport and the case study evaluation dataset. Evaluation methods described in the Evaluation Procedure section of Supplement.

### Outcomes

Automated assessments of reporting guideline adherence were compared against gold-standard human assessments using classification performance metrics including accuracy, precision, recall, and F1 score, the inter-assessor agreement metric Cohen’s kappa^32^, and operative metrics including cost and run-time reported as an mean average and 95% confidence interval. Mean classification performance and operative metrics were compared between different combinations of LLMs and prompting methods. For the SPIRIT-CONSORT-TM evaluation, performance metrics were reported at the article level for the full 83-element set and for the CONSORT-specific and SPIRIT-specific subsets reported in the SPIRIT-CONSORT-TM corpus. For the BenchReport evaluation, performance metrics were reported at the article level across all items of the original reporting guidelines with no modification. Outcome methods described in the Outcomes section of Supplement.

### Statistics

Summary measures of articleClevel means and associated 95 % CIs were reported. Differences between prompt methods and LLMs were compared using the twoCsided Wilcoxon test. To control multiplicity, we applied the Benjamini–Hochberg method for false discovery rate control at q=0.05 within each family of related hypotheses, defined as 1) multiple RAG methods for a fixed model and prompt method or 2) multiple LLMs for a fixed RAG method and prompt method.” Weighted mean accuracy was calculated by averaging review-level accuracies in proportion to the number of articles contributed by each review, so that reviews with more included studies contributed proportionally greater influence on the weighted mean accuracy estimate than reviews with fewer included studies. Statistical analyses were conducted using Python 3.8.9 and *scipy* 1.11.3. Statistical methods described in the Statistics section of Supplement.

### Ethics

This study did not require institutional review board (IRB) approval as it exclusively utilized publicly available, de-identified datasets. No human subjects were directly involved.

## Results

Through iterative testing of prompt engineering and RAG methods on the CONSORT Train set (n=10), we developed a final template prompt specifying the LLM’s role, guideline details, RAG-based article texts, and zero- or few-shot reporting examples (Figure 1A). On the CONSORT Validate set (n=20), we evaluated eight combinations of prompt design (zero-shot, few-shot) and RAG retrieval methods (no retrieval, contextual retrieval, BM25S retrieval, hybrid retrieval) with generalist (gpt-4o-mini) and reasoning (o3-mini) LLMs (Supplementary Table 3). For the reasoning LLM, no retrieval outperformed BM25S and hybrid retrieval but was comparable to contextual retrieval (Figure 1B), whereas the generalist LLM showed no differences in accuracy (Figure 1C). Notably, zero-shot prompting was non-inferior to few-shot for both generalist (Figure 1D) or reasoning (Figure 1E) LLMs. For the reasoning LLM, no retrieval had the highest cost but the lowest run time. For the generalist LLM, contextual and hybrid retrieval had the highest cost and run time than no retrieval or BM25S (Figure 1F). Overall, zero-shot with no retrieval was the best-performing engineered prompt and RAG combination based on classification performance and runtime, prompting further testing of the reasoning LLM with this prompt method as the AutoReporter tool.

**Figure 1.**
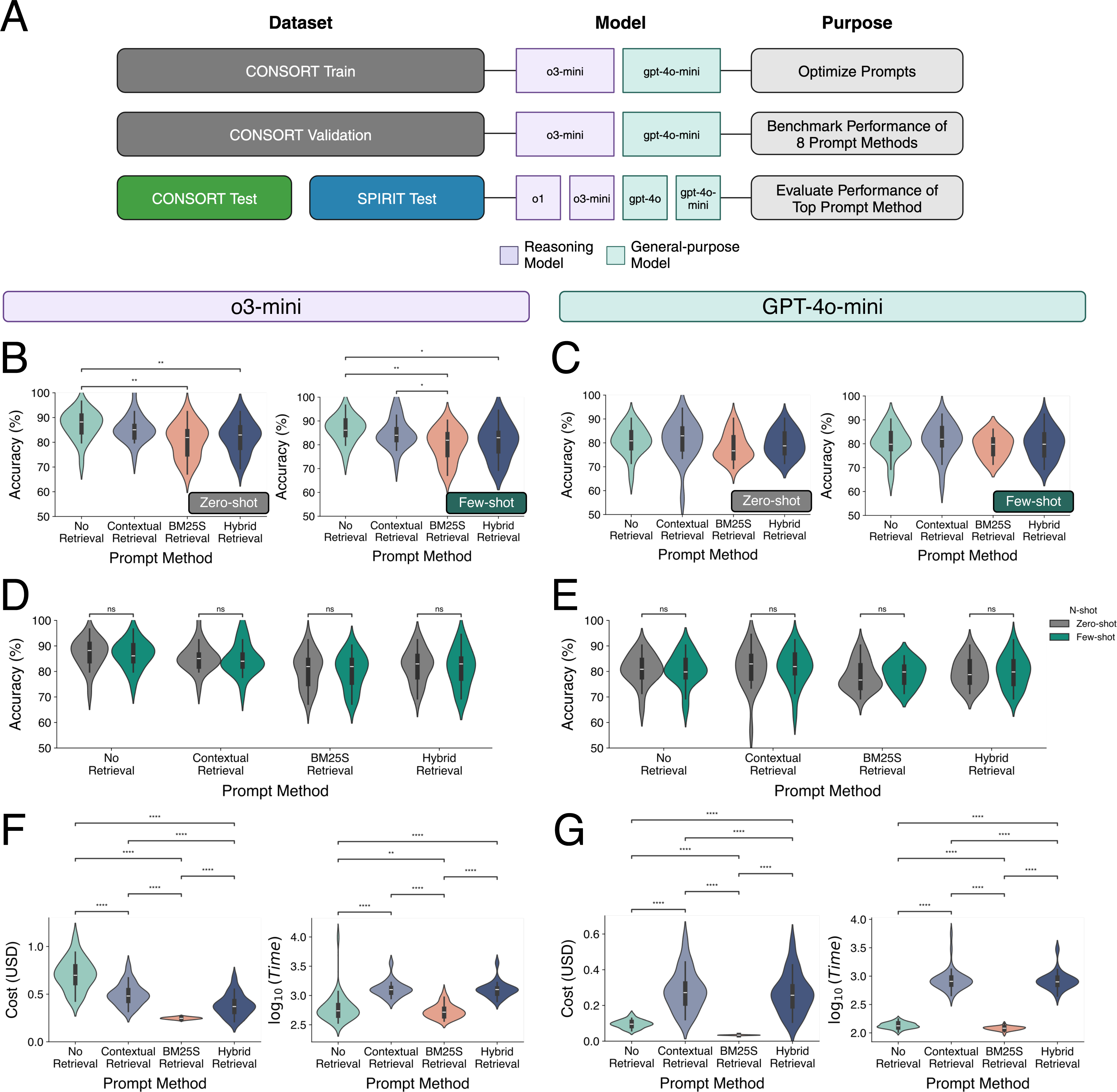
Evaluation of prompt methods applied to reasoning and general-purpose LLMs and tested on the CONSORT Validation dataset splits. A) Diagram of study workflow for initial evaluation of prompt methods applied to LLMs using the CONSORT Train, Validation, and Test dataset splits. B) Classification accuracy of the reasoning LLM (o3-mini) using zero-shot and few-shot prompting, stratified based on RAG method. C) Classification accuracy of the general-purpose LLM (gpt-4o-mini) using zero-shot and few-shot prompting, stratified based on RAG method. D) Classification accuracy of the reasoning LLM (o3-mini) using RAG methods, stratified based on zero- or few-shot prompting method. E) Classification accuracy of the reasoning LLM (gpt-4o-mini) using RAG methods, stratified based on zero- or few-shot prompting method. F) Cost and Time of the reasoning LLM (o3-mini) using zero-shot prompting, stratified based on RAG method. F) Cost and Time of the general-purpose LLM (gpt-4o-mini) using zero-shot prompting, stratified based on RAG method.

Evaluation of AutoReporter on the CONSORT Validate (n=20) and Test (n=20) datasets showed no differences in accuracy, precision, or recall between the generalist and reasoning LLMs, confirming generalizability on hold-out data (Supplementary Figure 3). Zero-shot remained non-inferior to few-shot across both the CONSORT (Supplementary Figure 4A, Supplementary Table 3) and SPIRIT (Supplementary Figure 4B, Supplementary Table 4) Test datasets, confirm our previous finding that zero-shot was non-inferior to few-shot prompting in the CONSORT Validate dataset. The reasoning o3-mini LLM achieved performance comparable to the o1 LLM and exceeded that of the generalist gpt-4o-mini and gpt-4o models in both CONSORT (Figure 2A) and SPIRIT Test evaluations (Figure 2B, Table 1). Performance and operational metrics on the CONSORT and SPIRIT Test datasets are provided in Supplementary Tables 4 and 5.

**Figure 2.**
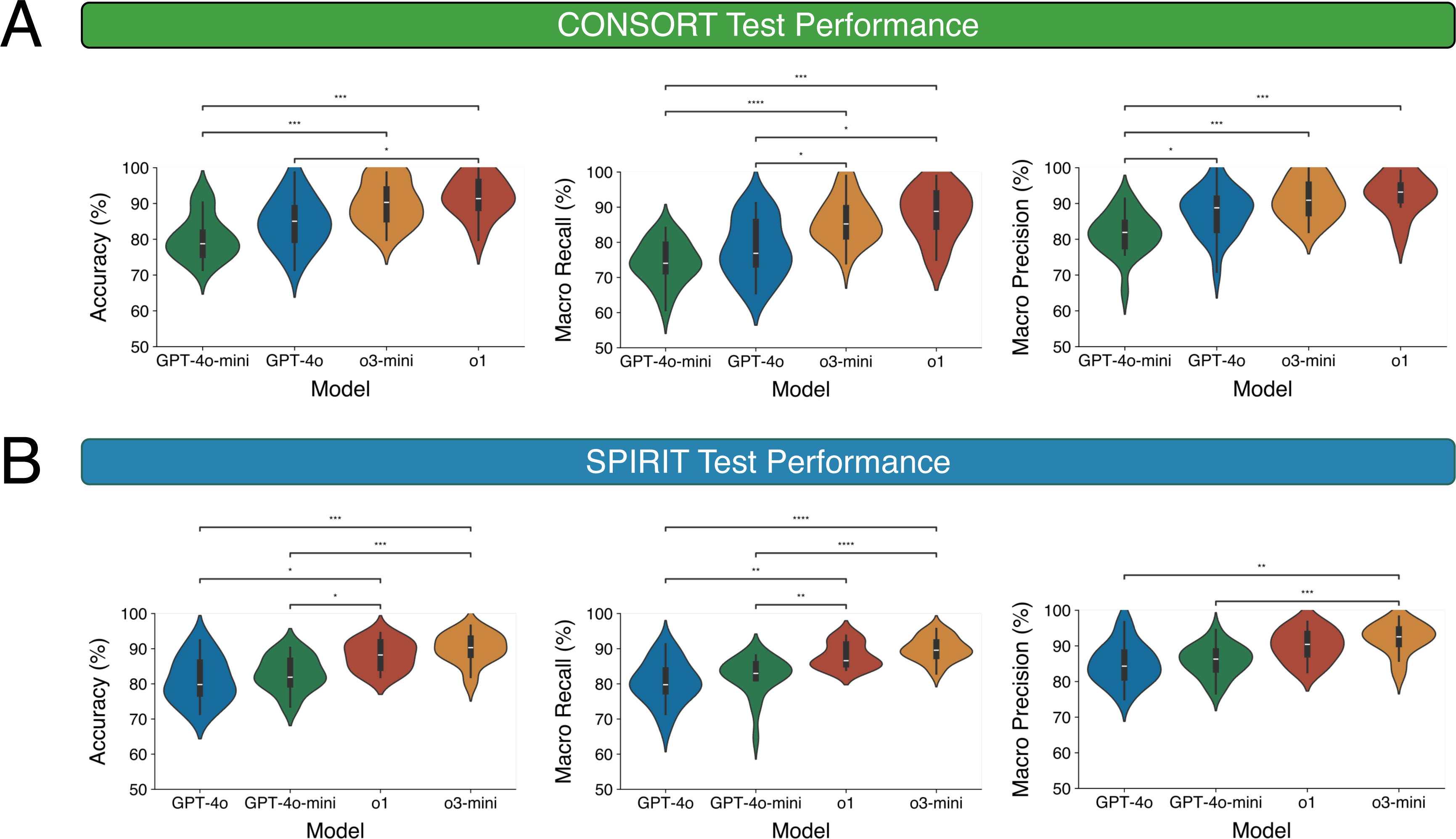
Evaluation of the top-performing zero-shot, no RAG retrieval prompt method tested on the CONSORT Test and SPIRIT Test dataset splits. A) Accuracy, macro recall, and macro precision of classification performance of the zero-shot, no RAG retrieval prompt method applied to reasoning (o1, o3-mini) and general-purpose (gpt-4o, gpt-4o-mini) LLMs and tested on the CONSORT Test dataset. A) Accuracy, macro recall, and macro precision of classification performance of the zero-shot, no RAG retrieval prompt method applied to reasoning (o1, o3-mini) and general-purpose (gpt-4o, gpt-4o-mini) LLMs and tested on the SPIRIT Test dataset.

**Table 1.** Classification and operative metrics of the zero-shot, no RAG retrieval prompt method applied to reasoning (o1, o3-mini) and general-purpose (gpt-4o, gpt-4o-mini) LLMs compared to the state-of-the-art PubMedBERT model using the CONSORT Test and SPIRIT Test dataset splits.

| Test Set | Metric | GPT-4o | GPT-4o-mini | o1 | o3-mini | SOTA PubMedBERT |
| --- | --- | --- | --- | --- | --- | --- |
| CONSORT | Accuracy | 84.63 (81.39 - 87.87) | 80.54 (77.68 - 83.39) | 91.65 (88.37 - 94.93) | 90.09 (87.20 - 92.98) | N.R. |
|  | Macro Precision | 87.63 (84.49 - 90.76) | 81.62 (78.85 - 84.38) | 92.74 (89.42 - 96.07) | 91.21 (88.63 - 93.79) | 87.5 |
|  | Micro Precision | 84.63 (81.39 - 87.87) | 80.54 (77.68 - 83.39) | 91.65 (88.37 - 94.93) | 90.09 (87.20 - 92.98) | 92.4 |
|  | Macro Recall | 78.27 (74.41 - 82.12) | 74.22 (71.41 - 77.04) | 88.12 (83.89 - 92.35) | 86.16 (83.16 - 89.16) | 85.9 |
|  | Micro Recall | 84.63 (81.39 - 87.87) | 80.54 (77.68 - 83.39) | 91.65 (88.37 - 94.93) | 90.09 (87.20 - 92.98) | 91.8 |
|  | Macro F1 | 79.24 (75.53 - 82.94) | 74.43 (71.63 - 77.23) | 89.31 (85.33 - 93.28) | 87.34 (84.43 - 90.25) | 85.8 |
|  | Micro F1 | 84.63 (81.39 - 87.87) | 80.54 (77.68 - 83.39) | 91.65 (88.37 - 94.93) | 90.09 (87.20 - 92.98) | 92.1 |
|  | Time (seconds) | 405.53 (318.38 - 492.68) | 349.82 (267.19 - 432.45) | 868.74 (725.50 - 1011.99) | 617.26 (499.52 - 735.00) | N.R. |
|  | Cost (USD) | 1.22 (1.06 - 1.38) | 0.31 (0.25 - 0.37) | 5.72 (4.77 - 6.68) | 0.68 (0.58 - 0.78) | N.R. |
| SPIRIT | Accuracy | 86.98 (85.02 - 88.93) | 83.45 (81.33 - 85.58) | 90.34 (87.91 - 92.76) | 92.07 (89.89 - 94.25) | N.R. |
|  | Macro Precision | 88.60 (86.77 - 90.43) | 84.30 (82.18 - 86.42) | 90.44 (88.10 - 92.77) | 92.85 (90.68 - 95.02) | 84.6 |
|  | Micro Precision | 86.98 (85.02 - 88.93) | 83.45 (81.33 - 85.58) | 90.34 (87.91 - 92.76) | 92.07 (89.89 - 94.25) | 91.7 |
|  | Macro Recall | 84.93 (83.00 - 86.86) | 81.34 (78.67 - 84.01) | 89.08 (86.66 - 91.50) | 90.84 (88.96 - 92.72) | 79.3 |
|  | Micro Recall | 86.98 (85.02 - 88.93) | 83.45 (81.33 - 85.58) | 90.34 (87.91 - 92.76) | 92.07 (89.89 - 94.25) | 87.1 |
|  | Macro F1 | 85.12 (83.26 - 86.99) | 81.12 (78.69 - 83.54) | 89.16 (86.65 - 91.66) | 91.08 (89.03 - 93.12) | 81 |
|  | Micro F1 | 86.98 (85.02 - 88.93) | 83.45 (81.33 - 85.58) | 90.34 (87.91 - 92.76) | 92.07 (89.89 - 94.25) | 89.4 |
|  | Time (seconds) | 391.35 (333.11 - 449.58) | 335.16 (285.16 - 385.16) | 884.15 (793.77 - 974.53) | 544.51 (484.52 - 604.50) | N.R. |
|  | Cost (USD) | 1.16 (0.91 - 1.42) | 0.31 (0.20 - 0.42) | 5.71 (4.18 - 7.24) | 0.65 (0.48 - 0.82) | N.R. |

The o3-mini LLM with zero-shot, no-retrieval prompts consistently matched or surpassed PubMedBERT on both CONSORT (92.7% vs. 87.5% macro precision) and SPIRIT (92.9% vs. 84.6% macro precision, 90.8% vs. 79.3% macro recall, 91.1% vs. 81.0% macro F1) Test evaluations (Table 1). The o3-mini LLM also performed comparably to the o1 LLM (90.09% vs. 91.65% accuracy) at substantially lower cost (US$0.68 vs. US$5.72). These findings informed the development of AutoReporter, which uses the top-performing, cost-effective o3-mini LLM with zero-shot, no-retrieval prompting for guideline adherence assessment.

To confirm generalizability beyond SPIRIT-CONSORT-TM, we created the public BenchReport dataset, composed of 10 systematic reviews where each systematic review assessed the item-level adherence of included studies to one research reporting guideline. BenchReport comprises a total of 506 article texts and 10,710 human ratings of reporting item adherence (Supplementary Table 5). AutoReporter achieved over 90% accuracy in 8 of 10 reviews and substantial agreement with human raters (Cohen’s κ>0.6) in 9 of 10 reviews (Table 2), with an overall weighted mean accuracy of 91.8%, macro precision of 90.0%, and macro recall of 91.9% (Figure 3).

**Figure 3.**
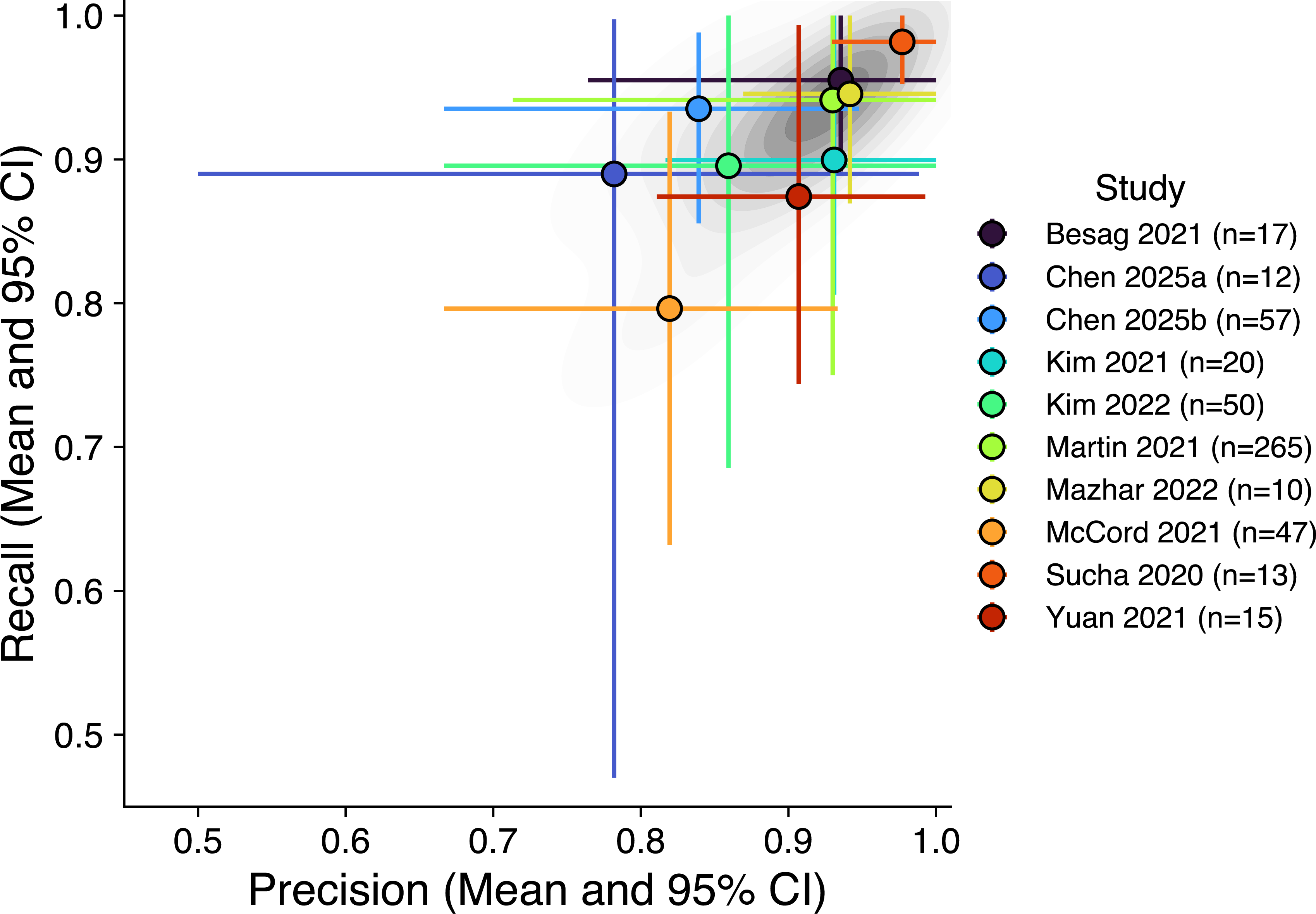
Mean and 95% of the macro precision and macro recall of AutoReporter, using the zero-shot, no RAG retrieval prompt method applied to the reasoning o3-mini LLM, evaluated on the BenchReport benchmark dataset.

**Table 2.**
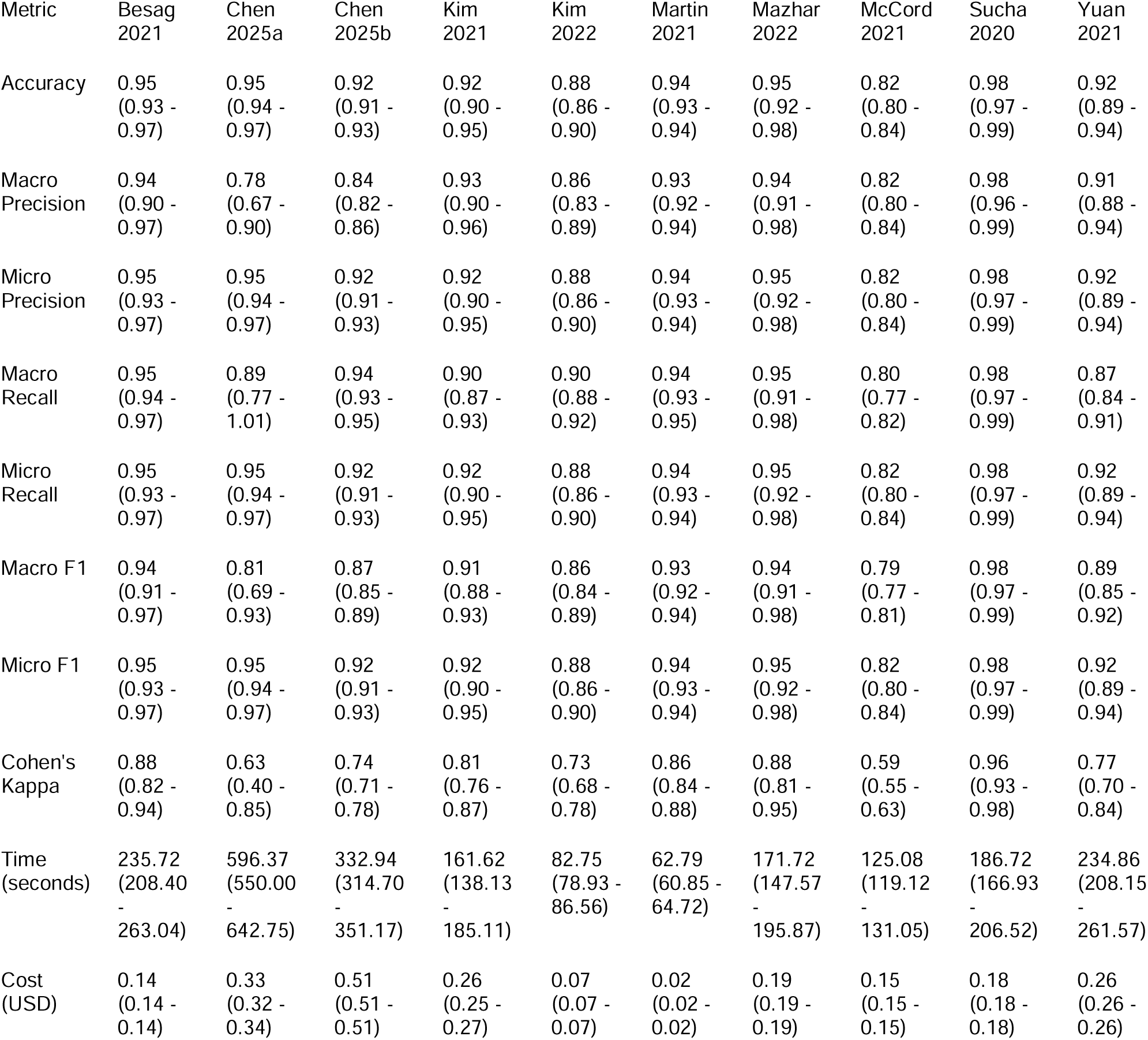
Classification and operative metrics of AutoReporter, the zero-shot, no RAG retrieval prompt method applied to the reasoning o3-mini LLM, across 10 systematic reviews in the BenchReport benchmark dataset.

| Metric | Besag<br>2021 | Chen<br>2025a | Chen<br>2025b | Kim<br>2021 | Kim<br>2022 | Martin<br>2021 | Mazhar<br>2022 | McCord<br>2021 | Sucha<br>2020 | Yuan<br>2021 |
| --- | --- | --- | --- | --- | --- | --- | --- | --- | --- | --- |
| Accuracy | 0.95<br>(0.93 -<br>0.97) | 0.95<br>(0.94 -<br>0.97) | 0.92<br>(0.91 -<br>0.93) | 0.92<br>(0.90 -<br>0.95) | 0.88<br>(0.86 -<br>0.90) | 0.94<br>(0.93 -<br>0.94) | 0.95<br>(0.92 -<br>0.98) | 0.82<br>(0.80 -<br>0.84) | 0.98<br>(0.97 -<br>0.99) | 0.92<br>(0.89 -<br>0.94) |
| Macro<br>Precision | 0.94<br>(0.90 -<br>0.97) | 0.78<br>(0.67 -<br>0.90) | 0.84<br>(0.82 -<br>0.86) | 0.93<br>(0.90 -<br>0.96) | 0.86<br>(0.83 -<br>0.89) | 0.93<br>(0.92 -<br>0.94) | 0.94<br>(0.91 -<br>0.98) | 0.82<br>(0.80 -<br>0.84) | 0.98<br>(0.96 -<br>0.99) | 0.91<br>(0.88 -<br>0.94) |
| Micro<br>Precision | 0.95<br>(0.93 -<br>0.97) | 0.95<br>(0.94 -<br>0.97) | 0.92<br>(0.91 -<br>0.93) | 0.92<br>(0.90 -<br>0.95) | 0.88<br>(0.86 -<br>0.90) | 0.94<br>(0.93 -<br>0.94) | 0.95<br>(0.92 -<br>0.98) | 0.82<br>(0.80 -<br>0.84) | 0.98<br>(0.97 -<br>0.99) | 0.92<br>(0.89 -<br>0.94) |
| Macro<br>Recall | 0.95<br>(0.94 -<br>0.97) | 0.89<br>(0.77 -<br>1.01) | 0.94<br>(0.93 -<br>0.95) | 0.90<br>(0.87 -<br>0.93) | 0.90<br>(0.88 -<br>0.92) | 0.94<br>(0.93 -<br>0.95) | 0.95<br>(0.91 -<br>0.98) | 0.80<br>(0.77 -<br>0.82) | 0.98<br>(0.97 -<br>0.99) | 0.87<br>(0.84 -<br>0.91) |
| Micro<br>Recall | 0.95<br>(0.93 -<br>0.97) | 0.95<br>(0.94 -<br>0.97) | 0.92<br>(0.91 -<br>0.93) | 0.92<br>(0.90 -<br>0.95) | 0.88<br>(0.86 -<br>0.90) | 0.94<br>(0.93 -<br>0.94) | 0.95<br>(0.92 -<br>0.98) | 0.82<br>(0.80 -<br>0.84) | 0.98<br>(0.97 -<br>0.99) | 0.92<br>(0.89 -<br>0.94) |
| Macro F1 | 0.94<br>(0.91 -<br>0.97) | 0.81<br>(0.69 -<br>0.93) | 0.87<br>(0.85 -<br>0.89) | 0.91<br>(0.88 -<br>0.93) | 0.86<br>(0.84 -<br>0.89) | 0.93<br>(0.92 -<br>0.94) | 0.94<br>(0.91 -<br>0.98) | 0.79<br>(0.77 -<br>0.81) | 0.98<br>(0.97 -<br>0.99) | 0.89<br>(0.85 -<br>0.92) |
| Micro F1 | 0.95<br>(0.93 -<br>0.97) | 0.95<br>(0.94 -<br>0.97) | 0.92<br>(0.91 -<br>0.93) | 0.92<br>(0.90 -<br>0.95) | 0.88<br>(0.86 -<br>0.90) | 0.94<br>(0.93 -<br>0.94) | 0.95<br>(0.92 -<br>0.98) | 0.82<br>(0.80 -<br>0.84) | 0.98<br>(0.97 -<br>0.99) | 0.92<br>(0.89 -<br>0.94) |
| Cohen's<br>Kappa | 0.88<br>(0.82 -<br>0.94) | 0.63<br>(0.40 -<br>0.85) | 0.74<br>(0.71 -<br>0.78) | 0.81<br>(0.76 -<br>0.87) | 0.73<br>(0.68 -<br>0.78) | 0.86<br>(0.84 -<br>0.88) | 0.88<br>(0.81 -<br>0.95) | 0.59<br>(0.55 -<br>0.63) | 0.96<br>(0.93 -<br>0.98) | 0.77<br>(0.70 -<br>0.84) |
| Time<br>(seconds) | 235.72<br>(208.40<br>-<br>263.04) | 596.37<br>(550.00<br>-<br>642.75) | 332.94<br>(314.70<br>-<br>351.17) | 161.62<br>(138.13<br>-<br>185.11) | 82.75<br>(78.93 -<br>86.56) | 62.79<br>(60.85 -<br>64.72) | 171.72<br>(147.57<br>-<br>195.87) | 125.08<br>(119.12<br>-<br>131.05) | 186.72<br>(166.93<br>-<br>206.52) | 234.86<br>(208.15<br>-<br>261.57) |
| Cost<br>(USD) | 0.14<br>(0.14 -<br>0.14) | 0.33<br>(0.32 -<br>0.34) | 0.51<br>(0.51 -<br>0.51) | 0.26<br>(0.25 -<br>0.27) | 0.07<br>(0.07 -<br>0.07) | 0.02<br>(0.02 -<br>0.02) | 0.19<br>(0.19 -<br>0.19) | 0.15<br>(0.15 -<br>0.15) | 0.18<br>(0.18 -<br>0.18) | 0.26<br>(0.26 -<br>0.26) |

We expect that users of AutoReporter may first assess an article draft for reporting guideline adherence, make revisions to the draft text to address reporting deficiencies, and then run AutoReporter’s assessment to confirm that indeed these revisions have wholly addressed the reporting deficiencies. To pilot this real-world use case, we sampled 10 systematic reviews with a mean PRISMA adherence of 73.10% (95% CI 62.72–83.47%) assessed by AutoReporter. In consultation with the primary or supervising author of each review (H.C.Y.W. [n=5], M.S.F.L [n=5]), we revised the article text based on the AutoReporter assessment to address item-specific reporting deficiencies. AutoReporter assessed all revised manuscripts to have 100% PRISMA adherence, confirming that the tool can effectively detect the edits made to address item-specific reporting deficiencies.

## Discussion

In this study aimed to design AutoReporter, an LLM-based tool for automated research reporting guideline assessment, we observed that 1) zero-shot with no RAG retrieval achieves superior performance compared to few-shot and RAG retrieval techniques, 2) the AutoReporter tool achieves non-inferior performance compared to the SOTA PubMedBERT method while forgoing the need for computational resources and annotated datasets for LLM fine-tuning, and 3) the AutoReporter tool generalizes well to assessing adherence across diverse sets of reporting guidelines, clinical domains, and article types.

The iterative design AutoReporter shows that a zero-shot, no RAG retrieval strategy paired with the reasoning o3-mini LLM delivers the optimal balance of classification performance, cost, and runtime. In both the CONSORT and SPIRIT test sets, our prompt strategy outperformed more complex fewCshot and RAG techniques and matched—or exceeded—the stateCofCtheCart PubMedBERT fineCtuned approach ^21^, while forgoing the need for large annotated data or resourceCintensive training. This finding aligns with reports of strong zero-shot reasoning on logical reasoning benchmarks ^33^ and clinical applications, such as patient trial matching ^34^ and diagnostic inference ^35^. We speculate that the superiority of the no RAG retrieval method is due to the long context windows of modern LLMs, obviating the additional RAG procedures of chunk selection and ranking ^36^. Notably, we observed that careful prompt structuring alone—role definition, context, and concise instructions—can optimize the latent reasoning in reasoning models without the need for resource-intensive, multi-step data retrieval for item-wise rating compared to prior methods ^19^, supporting previous observations of the utility of instruction structure optimization for biomedical systematic review screening ^37^.

Although AutoReporter tool was non-inferior to the state-of-the-art fine-tuned PubMedBERT ^21^ on the SPIRIT-CONSORT-TM corpus, we caution that domain-specific fine-tuning remains a viable approach to design LLMs for specialized NLP tasks. Prior work has shown that prompt engineering can out-perform fine-tuning for general medical question-answering ^38^, patient diagnosis classification ^39^, and clinical named-entity recognition ^40^ from clinical notes. Determining the cost-benefit analysis of fine-tuning compared to zero-shot prompting alone remains a caseCbyCcase decision. Applied to the BenchReport corpus, AutoReporter reliably achieved high classification performance across diverse reporting guidelines, clinical domains, and article types, underscoring the system’s generalizability in real-world biomedical literature. In the Bench-Report benchmark, reviews with fewer articles tended to have wider variability of AutoReporter performance, suggesting that performance estimates should be interpreted cautiously given that performance may be unstable and less generalizable to larger or more diverse corpora. Overall, we hypothesize that the underlying reasoning o3-mini LLM of AutoReporter may adapt to new contexts with minimal task-specific modification ^41^, enabling, in principle, broad generalizability across all guidelines, including evolving or newly introduced guidelines and guidelines in the EQUATOR network catalogue not assessed in this study. However, we caution that validation on emerging guideline and biomedical domains remains an important direction for future research.

Potential failure modes include missing important reporting details located in tables, figures, or appendices that may be overlooked when only narrative text is processed (Supplementary Table 6). Checklist items that are composite or ambiguously phrased may yield inconsistent outputs if the model exhibits mixed performance in assessing sub-component reporting and generating a consensus item-level reporting decision. Finally, semantically similar sentences within the same section could lead to mis-attributed quoted evidence used to support AutoReporter decisions. Future improvements could involve structured table and figure parsing, hierarchical item representation of items consisting of multiple sub-items, and self-verification methods to assess evidence attribution. We suggest that AutoReporter can function as an AI co-reviewer that flags missing items before peer review, mirroring the dual-reviewer schema in systematic reviews ^42^. Authors can iteratively refine manuscript drafts with explainable reporting assessments while journal editors can assess reporting adherence of submissions at scale.

Systematic performance evaluation of open-weight models and alternative providers of proprietary models merit evaluation as a future direction of research. Although this study reported accuracy for comparability with prior work, we acknowledge that accuracy can overstate performance in imbalanced tasks where common checklist items dominate, so we suggest that macro-averaged metrics and agreement methods can provide a more balanced view of performance and should be prioritized when interpreting AutoReporter’s performance. AutoReporter generated quoted evidence to support item reporting assessment and recommendations to improve adherence for non-reported items should be considered illustrative features and were not systematically validated, warranting future studies to assess whether these features provide accurate and actionable feedback in real-world manuscript preparation and review. There remains the possibility of data leakage where included texts appear in LLM pre-training corpora which may bias performance. Future evaluations should quantify whether prompt or article length, linguistic features, and language influences performance. The zeroCshot approach relies on the LLM’s implicit understanding of biomedical language, but fineCtuning or RAG may still improve performance in technical fields. Finally, prospective trials embedded in editorial workflows are needed to quantify user acceptance and the realCworld impact on manuscript quality.

## Conclusion

AutoReporter demonstrates that a zeroCshot reasoning LLM can output fast, lowCcost, and accurate assessments of reporting guideline adherence, outperforming both retrievalCaugmented and fineCtuned domain-specific models. AutoReporter’s strong performance across multiple guidelines and article types, without fineCtuning or retrieval, offers an automated, generalizable solution for authors, reviewers, and journal editors to assess the completeness of scientific research reporting.

## Data Availability

Data generated and used in this study is available at https://github.com/davidchen0420/AutoReporter-Data/tree/main.

## Code Availability

Code generated and used in this study is available at https://github.com/patrickli53/autoreporter. Open access version of the AutoReporter tool is available at https://autoreporter.streamlit.app.

## Acknowledgements

None.

