## Supplement for "AutoReporter: Development of an artificial intelligence tool for automated assessment of research reporting guideline adherence"

**Supplementary Methods**. Study methodology details.

**Supplementary Figure 1**. Diagram of SPIRIT-CONSORT-TM and BenchReport datasets used to evaluate the performance of the AutoReporter tool.

**Supplementary Figure 2**. Diagram of Contextual Retrieval RAG method applied to the AutoReporter tool.

**Supplementary Figure 3**. Comparison of classification performance of the zero-shot, no RAG retrieval prompt method applied to the reasoning (o3-mini) and generalist (gpt-4o-mini) LLM between the CONSORT Validation dataset and CONSORT Test dataset. A) Accuracy evaluation. B) Macro precision evaluation. C) Macro recall.

**Supplementary Figure 4**. Comparison of classification performance of the zero-shot, no RAG retrieval prompt method to the few-shot, no RAG retrieval prompt method applied to the reasoning (o1, o3-mini) and generalist (gpt-4o, gpt-4o-mini) LLM. A) CONSORT Test evaluation. B) SPIRIT Test evaluation.

**Supplementary Figure 5**. Evaluation of the top-performing zero-shot, no RAG retrieval prompt method tested on the CONSORT Test and SPIRIT Test dataset splits. A) Accuracy, macro recall, and macro precision of classification performance of the zero-shot, no RAG retrieval prompt method applied to reasoning (o1, o3-mini) and general-purpose (gpt-4o, gpt-4o-mini) LLMs and tested on the CONSORT Test dataset. A) Accuracy, macro recall, and macro precision of classification performance of the zero-shot, no RAG retrieval prompt method applied to reasoning (o1, o3-mini) and general-purpose (gpt-4o, gpt-4o-mini) LLMs and tested on the SPIRIT Test dataset.

**Supplementary Table 1**. Characteristics of the BenchReport dataset.

**Supplementary Table 2.** Example set of input prompts and expected outputs of AutoReporter.

**Supplementary Table 3**. Classification performance and operative metrics of the zero-shot, no RAG retrieval prompt method applied to the reasoning (o1, o3-mini) and generalist (gpt-4o, gpt-4o-mini) LLM on the CONSORT Validation dataset split from the SPIRIT-CONSORT-TM corpus. Metrics are shown for all 83 CONSORT and SPIRIT checklist items and the 79 CONSORT-specific checklist items reported in the SPIRIT-CONSORT-TM corpus.

**Supplementary Table 4**. Classification performance and operative metrics of the zero-shot, no RAG retrieval prompt method and few-shot, no RAG retrieval prompt method applied to the reasoning (o1, o3-mini) and generalist (gpt-4o, gpt-4o-mini) LLM on the CONSORT Test dataset split from the SPIRIT-CONSORT-TM corpus. Metrics are shown for all 83 CONSORT and SPIRIT checklist items and the 79 CONSORT-specific checklist items reported in the SPIRIT-CONSORT-TM corpus.

**Supplementary Table 5**. Classification performance and operative metrics of the zero-shot, no RAG retrieval prompt method and few-shot, no RAG retrieval prompt method applied to the reasoning (o1, o3-mini) and generalist (gpt-4o, gpt-4o-mini) LLM on the SPIRIT Test dataset split from the SPIRIT-CONSORT-TM corpus. Metrics are shown for all 83 CONSORT and SPIRIT checklist items and the 72 SPIRIT-specific checklist items reported in the SPIRIT-CONSORT-TM corpus.

**Supplementary Table 6**. Examples of failure modes observed by AutoReporter.

This supplementary material has been provided by the authors to give readers additional information about their work.

### Supplementary Methods

Datasets

The SPIRIT-CONSORT-TM corpus is a dataset of 100 English RCT reports and 100 English RCT protocols with human annotations of whether 83 items were reported from SPIRIT and CONSORT checklists [21] (Supplementary Figure 1). We accessed the SPIRIT-CONSORT-TM corpus last updated on February 1, 2025. All dataset splits of the SPIRIT-CONSORT-TM used in this study were analyzed on March 5, 2025 and reported at https://github.com/davidchen0420/AutoReporter-Data/tree/main. No further pre-processing or handling of missing data of the SPIRIT-CONSORT-TM corpus was conducted.

The BenchReport corpus is a novel, public benchmark dataset generated on April 6, 2025 that consists of 8 randomly sampled published systematic reviews [22–29] and 2 unpublished systematic reviews of reporting guideline adherence that represents 10 reporting guidelines, 7 clinical domains, and 6 article types (Supplementary Table 1, Supplementary Figure 1). BenchReport includes a total of 10,710 human ratings of reporting guideline items for 506 English article texts. To design BenchReport, for each of the 10 systematic reviews, we collected the human binary rating of reporting guideline adherence from the review and sourced the full text of the articles assessed in the review. Full-text articles that were non-English or could not be accessed were excluded from BenchReport (n=20, shown in Supplementary Table 1). No further pre-processing or handling of missing data of the BenchReport corpus was conducted. The BenchReport used in this study was analyzed on April 10, 2025 and reported at https://github.com/davidchen0420/AutoReporter-Data/tree/main.

The case study evaluation of AutoReporter to identify edits made to address PRISMA reporting deficiencies included 10 randomly sampled systematic reviews first-authored or supervised by H.C.Y.W. (n=5) or M.S.F.L (n=5). This case study dataset was analyzed on April 15, 2025 and found at https://github.com/davidchen0420/AutoReporter-Data/tree/main.

LLM Design

We evaluated two generalist (gpt-4o, gpt-4o-mini) [30] and two reasoning (gpt-o1, gpt-o3-mini) [31]LLMs in this study, first accessed on March 5, 2025 and last accessed on April 15, 2025 using the OpenAI application programming interface. All LLMs were evaluated using default parameters with no change to seed, temperature, max token length, or penalties. Each LLM was prompted with an instruction that defined their role, to assess reporting guideline adherence, and a prompt, to assess adherence to each item in a reporting guideline. The input of the LLM included the article text that varied based on the RAG method used, the guideline item number, the guideline item description, examples of wholly reported guideline items depending on whether zero- or few-shot prompting was used, and the instructive prompt to assess guideline adherence. The output of the LLM included the binary rating of item reporting (0: not wholly reported; 1: wholly reported) and quoted evidence from the input text that supports the rating decision. For items that were not wholly reported, a recommendation was provided on how to revise the input text to address the item-specific reporting deficiency.

Prompt Method Design

Few-shot prompting included examples of the wholly reported guideline item in the LLM instruction prompt. For few-shot prompting tested on the SPIRIT-CONSORT-TM corpus, examples were sourced from the SPIRIT-CONSORT-TM annotation guide last updated on February 1, 2025 (https://osf.io/ha73p). Zero-shot prompting did not include any reference examples in the LLM instruction prompt.

We implemented the Contextual Retrieval RAG method first described by Anthropic on September 19, 2024 (https://www.anthropic.com/news/contextual-retrieval, Supplementary Figure 2). To pre-process input texts for RAG retrieval, for the full text of each given article, we chunked the text using a chunk size of 500 characters and overlap of 100 characters. For each text chunk, we prompted a generalist LLM (gpt-4o-mini) to generate a contextual description of the chunk situated within the overall input article, for the purpose of improving search retrieval of the chunk. For each generated chunk context appended to its source chunk, we generated a vector embedding of the text chunk and its associated context using an embedding LLM (text-embedding-3-large).

No RAG retrieval used the entire full-text article as input into the LLM for reporting guideline assessment. BM25S Retrieval is a lexical search RAG technique that ranks the text chunks and their contexts sourced from a full-text article and outputs the top 10 text chunks that are most relevant to the guideline item and description based on the co-occurence of shared terms. Contextual Retrieval is a context-based RAG technique that ranks the similarity of the vector embeddings of each text chunk and their context sourced from a full-text article to the vector embedding of the guideline item description. Contextual Retrieval outputs the top 10 text chunks that are most relevant to the guideline item and description based on rank-ordered cosine similarity of the vector embeddings of the text chunks and guideline item description. Hybrid Retrieval, also known as reciprocal rank fusion, is a rank aggregation method that combines rankings of retrieved text chunks by the BM25S Retrieval and Contextual Retrieval RAG methods. Hybrid Retrieval outputs the top 5 text chunks that were individually ranked as relevant to the guideline item and description by the BM25S Retrieval and Contextual Retrieval RAG methods.

Evaluation Procedure

The SPIRIT-CONSORT-TM Validate dataset was used to determining the top performing prompt method out of 8 total combinations. Two types of prompt engineering with examples (zero-shot, few-shot) and four types of RAG to retrieve relevant article text (no RAG retrieval, contextual retrieval, BM25S retrieval, hybrid retrieval) methods were tested, totaling 8 possible combinations of prompt engineering and RAG methods. We evaluated the 8 combinations of prompt methods applied to a generalist (gpt-4o-mini) and reasoning (gpt-o3-mini) LLM. The SPIRIT-CONSORT-TM Test dataset was used to test the classification performance of the top-performing zero-shot, no RAG retrieval prompt methodology using two generalist (gpt-4o, gpt-4o-mini) and two reasoning (gpt-o1, gpt-o3-mini) LLMs in comparison to the SOTA PubMedBERT model on the same dataset.

In the testing phase, we optimized template prompts using the CONSORT Train split (n=10) with generalist (gpt-4o-mini) and reasoning (gpt-o3-mini) LLMs. We tested the inclusion and ordering of defining the LLM's role, guideline item details, article text, examples of item reporting, and instructions to assess guideline adherence. Each iteration empirically assessed LLM adherence to reporting guidelines against human expert evaluations and reviewed the LLM's rationale to progressively improve output quality. In the validation phase, 8 pairwise combinations of prompt engineering with examples of reported CONSORT examples (zero-shot, few-shot) and retrieval augmented generation (RAG) to retrieve relevant article text (no retrieval, contextual retrieval, BM25S retrieval, hybrid retrieval) methods applied using either a generalist (gpt-4o-mini) or reasoning (gpt-o3-mini) LLM using the CONSORT Validate split (n=20). In the testing phase, we assessed the top-performing zero-shot, no RAG retrieval prompt method across generalist (gpt-4o, gpt-4o-mini) and reasoning (gpt-o1, gpt-o3-mini) LLMs using the CONSORT Test split (n=20) and SPIRIT Test split (n=20). The CONSORT and SPIRIT Test splits were the same dataset splits used by the state-of-the-art PubMedBERT method to enable direct performance comparisons.

The BenchReport dataset was used to test the classification performance of the top-performing zero-shot, no RAG retrieval prompt methodology using the top-performing, cost-effective reasoning LLM (gpt-o3-mini) across diverse biomedical texts that vary based on reporting guideline assessed, included article types, and clinical domain applications. The case study evaluation of AutoReporter was used to test the ability for AutoReporter to sensitively detect manual edits made by authors and guided by recommendations from AutoReporter to address reporting deficiencies identified by AutoReporter. For all evaluation scenarios, the binary LLM rating of item reporting was compared to the gold-standard binary human rating of item reporting for each article.

### Outcomes

To assess item-wise reporting guideline assessment performance compared to gold-standard human assessments, we used the classification performance metrics of accuracy, precision, recall, and F1 score as well as the inter-assessor agreement metric of Cohen's kappa [32]. Both macro and micro precision, recall, and F1 score were reported. Macro metrics offer insights into classification performance across all classes equally, highlighting the prediction robustness on underrepresented classes, while micro metrics reflect overall classification performance weighted by class frequency, emphasizing performance on majority classes. Cost and time of each assessment was used to characterize operative metrics related to LLM-based guideline assessment. For each SPIRIT-CONSORT-TM dataset split and each of the 10 reviews in the BenchReport corpus, we prompted AutoReporter to generate item-wise guideline assessmentsfor each article, and classification and inter-assessor agreement metrics were calculated and averaged across all articles in the dataset to determine a mean score of guideline assessment performance. Mean classification performance and operative metrics were compared between different combinations of LLMs and prompting methods.

### Statistics

The summary measures of the performance and operative metrics were calculated by averaging article-level values across all articles in the dataset to derive mean values and the t-distribution of the set of article-level values to generate a 95% confidence interval. Study-count weighted mean average performance metrics were used to measure overall classification performance on the BenchReport corpus. Using the Wilcoxon test with Benjamini-Hochberg multiple testing correction, we compared the means of the performance and operative metrics between different LLMs and prompt methods. Statistical analyses were conducted with Python 3.8.9 and scipy 1.11.3.

### Ethics

This study did not require institutional review board (IRB) approval as it exclusively utilized publicly available, de-identified datasets. No human subjects were directly involved.

**Supplementary Figure 1**. Diagram of SPIRIT-CONSORT-TM and BenchReport datasets used to evaluate the performance of the AutoReporter tool.

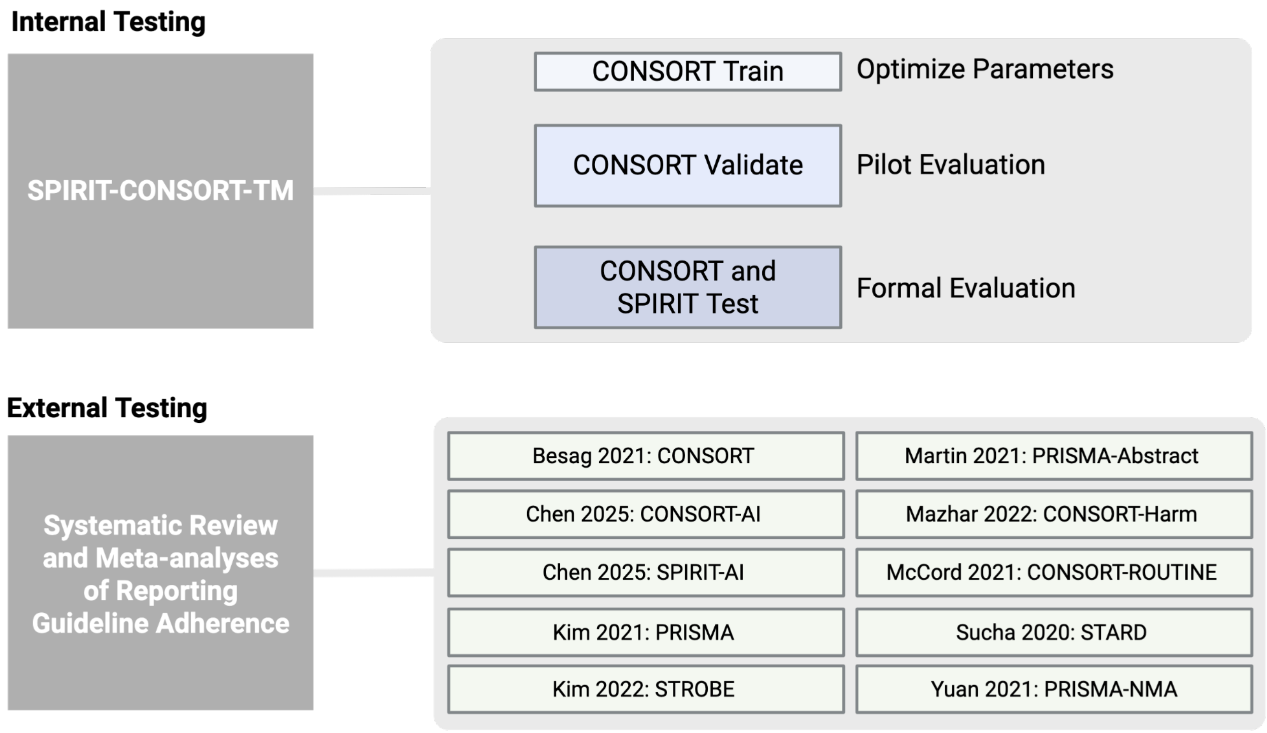

**Supplementary Figure 2**. Diagram of Contextual Retrieval RAG method applied to the AutoReporter tool. Full-text articles are chunked and augmented with an LLM-generated context that summarizes the chunk text. For each reporting guideline item, contextual retrieval ranks chunks and their context based on relevance to the item description. The top 5 most relevant chunks are then input to AutoReporter for reporting guideline adherence assessment.

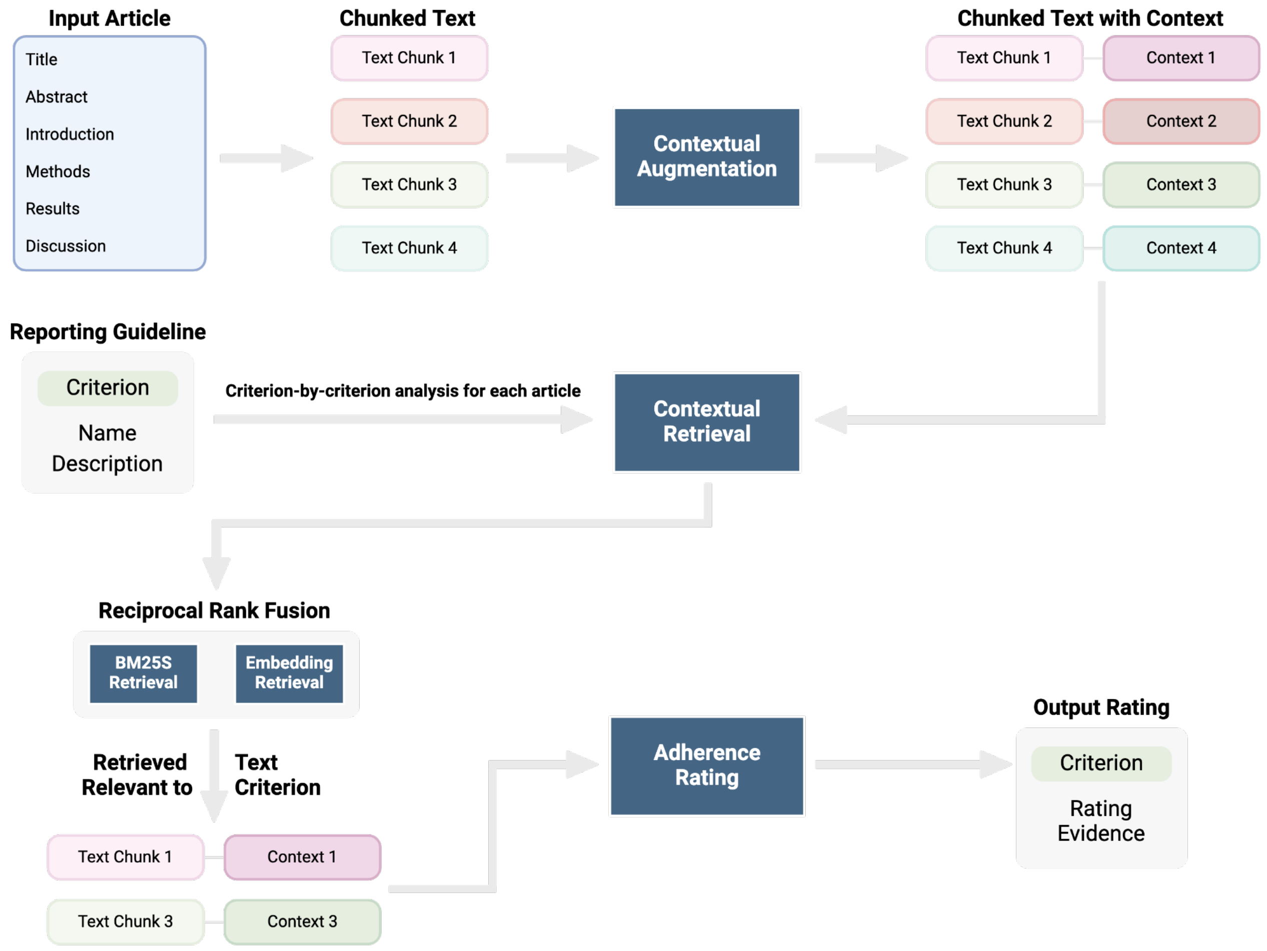

**Supplementary Figure 3**. Comparison of classification performance of the zero-shot, no RAG retrieval prompt method applied to the reasoning (o3-mini) and generalist (gpt-4o-mini) LLM between the CONSORT Validation dataset and CONSORT Test dataset. A) Accuracy evaluation. B) Macro precision evaluation. C) Macro recall evaluation.

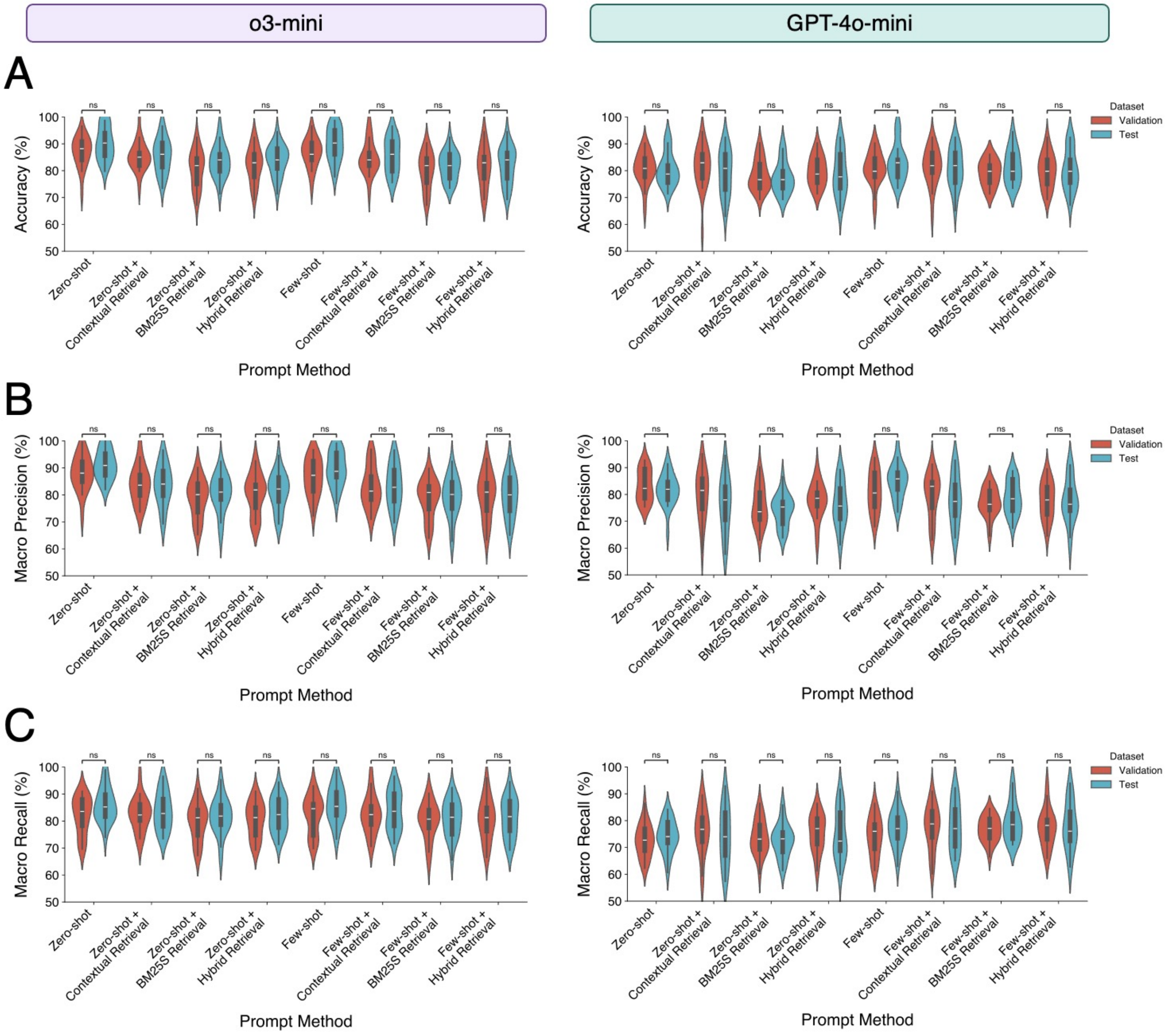

**Supplementary Figure 4**. Comparison of classification performance of the zero-shot, no RAG retrieval prompt method to the few-shot, no RAG retrieval prompt method applied to the reasoning (o1, o3-mini) and generalist (gpt-4o, gpt-4o-mini) LLM. A) CONSORT Test evaluation. B) SPIRIT Test evaluation.

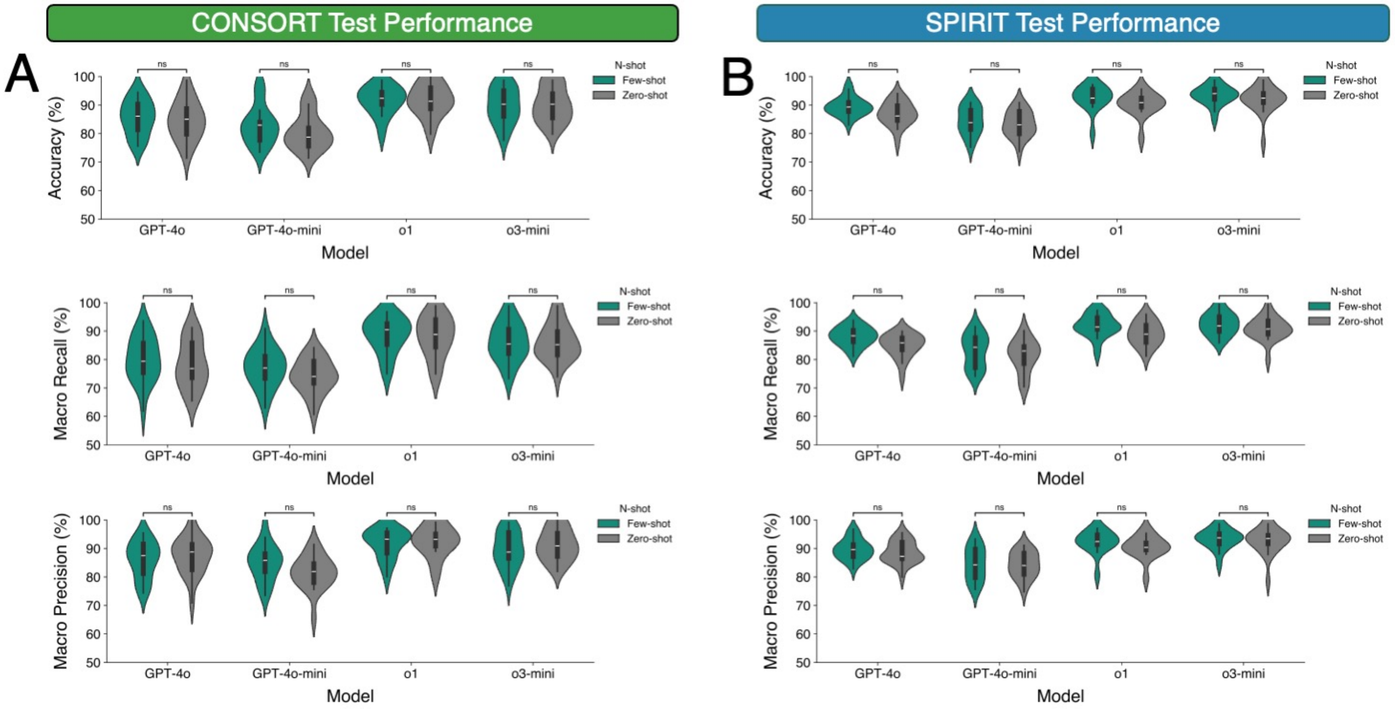

**Supplementary Figure 5**. Evaluation of the top-performing zero-shot, no RAG retrieval prompt method tested on the CONSORT Test and SPIRIT Test dataset splits. A) Accuracy, macro recall, and macro precision of classification performance of the zero-shot, no RAG retrieval prompt method applied to reasoning (o1, o3-mini) and general-purpose (gpt-4o, gpt-4o-mini) LLMs and tested on the CONSORT Test dataset. A) Accuracy, macro recall, and macro precision of classification performance of the zero-shot, no RAG retrieval prompt method applied to reasoning (o1, o3-mini) and general-purpose (gpt-4o, gpt-4o-mini) LLMs and tested on the SPIRIT Test dataset.

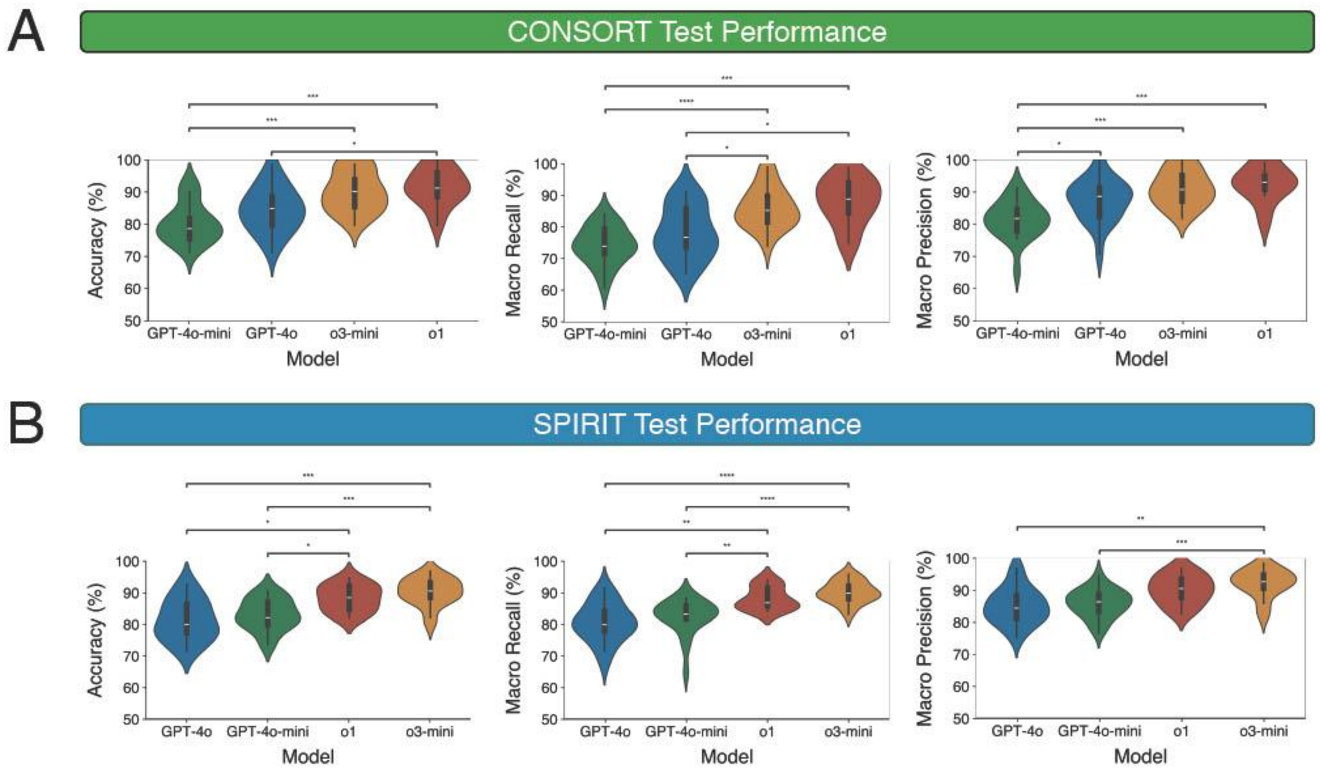

**Supplementary Table 1**. Characteristics of the BenchReport dataset. Full-text articles included in reviews that were non-English or could not be accessed were excluded from BenchReport.

| Study ID | PMID | Domain | Text Type | Number of Articles Included in Review | Number of Articles Included in BenchReport | Reporting Guideline | Reporting Guideline Number of Items |
|---|---|---|---|---|---|---|---|
| Besag 2021 | 34286606 | Psychiatry | Randomized Controlled Trial Report | 17 | 17 | CONSORT | 37 |
| Chen 2025a | Unpublished | Oncology | Randomized Controlled Trial Report | 57 | 57 | CONSORT-AI | 51 |
| Chen 2025b | Unpublished | Oncology | Randomized Controlled Trial Report | 12 | 12 | SPIRIT-AI | 66 |
| Kim 2021 | 34838030 | General | Systematic Review | 20 | 20 | PRISMA | 27 |
| Kim 2022 | 35615091 | Complementary and Alternative Medicine | Observational Study Report | 62 | 50 | STROBE | 12 |
| Martin 2021 | 33573591 | Dentistry | Abstract | 265 | 265 | PRISMA-Abstract | 12 |
| Mazhar 2022 | 33007453 | Infectious Disease | Trial Report | 16 | 10 | CONSORT-Harm | 20 |
| McCord 2021 | 34525408 | General | Randomized Controlled Trial Report | 47 | 47 | CONSORT-Routine | 20 |
| Sucha 2020 | 33778613 | Radiology | Trial Report | 13 | 13 | STARD | 34 |
| Yuan 2021 | 33505490 | Complementary and Alternative Medicine | Network Meta Analysis Report | 17 | 15 | PRISMA-NMA | 32 |

**Supplementary Table 2.** Example set of input prompts and expected outputs of AutoReporter.

| Input or Output | Example |
|---|---|
| Input: Role Instruction Prompt | As a research methods scientist, your role is to evaluate the adherence of full-text scientific research articles to established reporting guidelines. For each reporting item in the guideline, you will be assessing the research article to confirm if the item has been wholly reported or not. Your assessment approach should be adaptable to any reporting guideline and research domain. |
| Input: Example Task Instruction Prompt | ### Full Text of Article<br><br>[ ARTICLE TEXT ]<br><br>### Checklist Item<br><br>7<br><br>### Checklist Item Section<br><br>Methods<br><br>### Checklist Item Description<br><br>Present the full search strategies for all databases, registers and websites, including any filters and limits used.<br><br>### Task<br><br>Based on the provided article text, checklist item name, checklist item description, and checklist item section, is the checklist item reported in the full text of this article?<br><br>Think step-by-step. Be evidence-based and explicitly adherent to all components of the checklist item description when assessing checklist item reporting. |
| Output: Example Rating | 0 (Not Fully Reported) |
| Output: Example Evidence | The article states that a literature search was performed using Ovid MEDLINE, Embase, and Cochrane databases between January 1, 1946, and January 31, 2023, and mentions that the literature search methodology will be described in detail in a separate publication. However, it does not present the full search strategies, including all filters and limits used. |
| Output: Example Recommended Edit to Improve Reporting | To fully report the search strategy, include the complete search strings for each database along with any filters, limits, or specific search terms applied. |

**Supplementary Table 3**. Classification performance and operative metrics of the zero-shot, no RAG retrieval prompt method applied to the reasoning (o1, o3-mini) and generalist (gpt-4o, gpt-4o-mini) LLM on the CONSORT Validation dataset split from the SPIRIT-CONSORT-TM corpus. Metrics are shown for all 83 CONSORT and SPIRIT checklist items and the 79 CONSORT-specific checklist items reported in the SPIRIT-CONSORT-TM corpus.

| Model | Metric | Zero-shot All Item | Zero-shot + Contextual Retrieval All Item | Zero-shot + BM25S Retrieval All Item | Zero-shot + Hybrid Retrieval All Item | Few-shot All Item | Few-shot + Contextual Retrieval All Item | Few-shot + BM25S Retrieval All Item | Few-shot + Hybrid Retrieval All Item | Zero-shot CONSORT Item | Zero-shot + Contextual Retrieval CONSORT Item | Zero-shot + BM25S Retrieval CONSORT Item | Zero-shot + Hybrid Retrieval CONSORT Item | Few-shot CONSORT Item | Few-shot + Contextual Retrieval CONSORT Item | Few-shot + BM25S Retrieval CONSORT Item | Few-shot + Hybrid Retrieval CONSORT Item |
|---|---|---|---|---|---|---|---|---|---|---|---|---|---|---|---|---|---|
| GPT-4o-mini | Accuracy | 78.307 (76.160 - 80.454) | 78.623 (75.572 - 81.675) | 77.801 (76.025 - 79.577) | 78.244 (76.645 - 79.843) | 79.762 (77.543 - 81.981) | 81.470 (79.130 - 83.810) | 80.521 (78.522 - 82.520) | 80.837 (78.708 - 82.967) | 80.850 (78.061 - 83.639) | 81.165 (77.322 - 85.008) | 78.330 (75.641 - 81.019) | 79.800 (77.209 - 82.391) | 80.850 (77.936 - 83.764) | 81.480 (78.203 - 84.757) | 79.170 (76.913 - 81.427) | 80.010 (77.055 - 82.965) |
| GPT-o3-mini | Accuracy | 86.973 (84.621 - 89.325) | 85.202 (82.839 - 87.564) | 82.925 (80.212 - 85.638) | 83.620 (80.898 - 86.343) | 87.416 (85.312 - 89.519) | 85.139 (82.762 - 87.515) | 82.672 (79.967 - 85.377) | 83.873 (81.474 - 86.273) | 87.045 (84.330 - 89.760) | 85.575 (82.948 - 88.202) | 80.640 (77.413 - 83.867) | 82.215 (79.202 - 85.228) | 86.625 (84.057 - 89.193) | 84.735 (81.843 - 87.627) | 80.010 (76.823 - 83.197) | 81.375 (77.869 - 84.881) |
| GPT-4o-mini | Macro F1 | 76.283 (73.870 - 78.696) | 77.362 (74.156 - 80.567) | 76.835 (74.823 - 78.847) | 77.226 (75.323 - 79.128) | 78.392 (75.853 - 80.930) | 80.710 (78.170 - 83.251) | 80.051 (77.915 - 82.187) | 80.289 (78.019 - 82.558) | 73.522 (70.215 - 76.829) | 76.642 (72.228 - 81.056) | 74.103 (70.877 - 77.329) | 75.673 (72.443 - 78.903) | 74.732 (70.920 - 78.544) | 77.591 (73.669 - 81.513) | 76.284 (73.916 - 78.652) | 77.045 (73.897 - 80.194) |
| GPT-o3-mini | Macro F1 | 86.317 (83.736 - 88.897) | 84.697 (82.090 - 87.304) | 82.609 (79.834 - 85.385) | 83.228 (80.337 - 86.119) | 86.894 (84.561 - 89.227) | 84.744 (82.291 - 87.197) | 82.366 (79.602 - 85.130) | 83.537 (80.973 - 86.101) | 83.304 (79.931 - 86.677) | 82.841 (79.766 - 85.915) | 78.550 (75.144 - 81.957) | 79.881 (76.610 - 83.152) | 83.053 (79.645 - 86.462) | 82.138 (78.809 - 85.466) | 78.201 (74.747 - 81.655) | 79.406 (75.623 - 83.189) |
| GPT-4o-mini | Micro F1 | 78.307 (76.160 - 80.454) | 78.623 (75.572 - 81.675) | 77.801 (76.025 - 79.577) | 78.244 (76.645 - 79.843) | 79.762 (77.543 - 81.981) | 81.470 (79.130 - 83.810) | 80.521 (78.522 - 82.520) | 80.837 (78.708 - 82.967) | 80.850 (78.061 - 83.639) | 81.165 (77.322 - 85.008) | 78.330 (75.641 - 81.019) | 79.800 (77.209 - 82.391) | 80.850 (77.936 - 83.764) | 81.480 (78.203 - 84.757) | 79.170 (76.913 - 81.427) | 80.010 (77.055 - 82.965) |
| GPT-o3-mini | Micro F1 | 86.973 (84.621 - 89.325) | 85.202 (82.839 - 87.564) | 82.925 (80.212 - 85.638) | 83.620 (80.898 - 86.343) | 87.416 (85.312 - 89.519) | 85.139 (82.762 - 87.515) | 82.672 (79.967 - 85.377) | 83.873 (81.474 - 86.273) | 87.045 (84.330 - 89.760) | 85.575 (82.948 - 88.202) | 80.640 (77.413 - 83.867) | 82.215 (79.202 - 85.228) | 86.625 (84.057 - 89.193) | 84.735 (81.843 - 87.627) | 80.010 (76.823 - 83.197) | 81.375 (77.869 - 84.881) |
| GPT-4o-mini | Macro Precision | 83.443 (81.915 - 84.971) | 81.327 (78.509 - 84.145) | 79.141 (77.049 - 81.233) | 79.814 (78.341 - 81.287) | 82.855 (80.835 - 84.874) | 82.583 (80.338 - 84.828) | 80.740 (78.588 - 82.892) | 81.180 (79.058 - 83.302) | 83.300 (80.516 - 86.084) | 79.775 (75.412 - 84.137) | 75.754 (72.316 - 79.192) | 77.639 (74.507 - 80.771) | 81.130 (77.559 - 84.702) | 79.519 (75.673 - 83.364) | 77.043 (74.559 - 79.528) | 77.805 (74.577 - 81.032) |
| GPT-o3-mini | Macro Precision | 88.668 (86.874 - 90.462) | 85.566 (82.976 - 88.156) | 83.022 (80.321 - 85.722) | 83.582 (80.727 - 86.436) | 88.543 (86.672 - 90.414) | 85.348 (82.916 - 87.780) | 82.944 (80.139 - 85.749) | 83.792 (81.224 - 86.359) | 88.030 (84.978 - 91.081) | 84.255 (80.875 - 87.636) | 78.779 (75.459 - 82.099) | 80.270 (76.814 - 83.726) | 86.580 (83.357 - 89.804) | 83.056 (79.558 - 86.554) | 78.508 (75.154 - 81.862) | 79.828 (75.958 - 83.697) |
| GPT-4o-mini | Micro Precision | 78.307 (76.160 - 80.454) | 78.623 (75.572 - 81.675) | 77.801 (76.025 - 79.577) | 78.244 (76.645 - 79.843) | 79.762 (77.543 - 81.981) | 81.470 (79.130 - 83.810) | 80.521 (78.522 - 82.520) | 80.837 (78.708 - 82.967) | 80.850 (78.061 - 83.639) | 81.165 (77.322 - 85.008) | 78.330 (75.641 - 81.019) | 79.800 (77.209 - 82.391) | 80.850 (77.936 - 83.764) | 81.480 (78.203 - 84.757) | 79.170 (76.913 - 81.427) | 80.010 (77.055 - 82.965) |

| | | | | | | | | | | | | | | | | | |
|---|---|---|---|---|---|---|---|---|---|---|---|---|---|---|---|---|---|
| GPT-o3-mini | Micro Precision | 86.973 (84.621 - 89.325) | 85.202 (82.839 - 87.564) | 82.925 (80.212 - 85.638) | 83.620 (80.898 - 86.343) | 87.416 (85.312 - 89.519) | 85.139 (82.762 - 87.515) | 82.672 (79.967 - 85.377) | 83.873 (81.474 - 86.273) | 87.045 (84.330 - 89.760) | 85.575 (82.948 - 88.202) | 80.640 (77.413 - 83.867) | 82.215 (79.202 - 85.228) | 86.625 (84.057 - 89.193) | 84.735 (81.843 - 87.627) | 80.010 (76.823 - 83.197) | 81.375 (77.869 - 84.881) |
| GPT-4o-mini | Macro Recall | 77.496 (75.427 - 79.564) | 78.192 (75.340 - 81.045) | 77.385 (75.385 - 79.386) | 77.885 (76.121 - 79.649) | 79.220 (76.951 - 81.489) | 81.140 (78.724 - 83.556) | 80.361 (78.290 - 82.432) | 80.615 (78.372 - 82.858) | 72.645 (69.773 - 75.517) | 76.154 (72.064 - 80.245) | 73.891 (70.841 - 76.941) | 75.541 (72.305 - 78.776) | 74.117 (70.708 - 77.527) | 77.353 (73.520 - 81.186) | 76.783 (74.600 - 78.966) | 77.413 (74.127 - 80.698) |
| GPT-o3-mini | Macro Recall | 86.714 (84.272 - 89.156) | 84.925 (82.309 - 87.540) | 82.962 (80.287 - 85.636) | 83.473 (80.634 - 86.312) | 87.291 (85.002 - 89.580) | 84.957 (82.534 - 87.381) | 82.783 (80.123 - 85.443) | 83.830 (81.301 - 86.358) | 82.177 (78.934 - 85.420) | 82.633 (79.685 - 85.581) | 79.723 (76.484 - 82.962) | 80.504 (77.450 - 83.559) | 82.304 (78.988 - 85.619) | 82.351 (79.069 - 85.633) | 79.983 (76.766 - 83.200) | 80.640 (77.012 - 84.268) |
| GPT-4o-mini | Micro Recall | 78.307 (76.160 - 80.454) | 78.623 (75.572 - 81.675) | 77.801 (76.025 - 79.577) | 78.244 (76.645 - 79.843) | 79.762 (77.543 - 81.981) | 81.470 (79.130 - 83.810) | 80.521 (78.522 - 82.520) | 80.837 (78.708 - 82.967) | 80.850 (78.061 - 83.639) | 81.165 (77.322 - 85.008) | 78.330 (75.641 - 81.019) | 79.800 (77.209 - 82.391) | 80.850 (77.936 - 83.764) | 81.480 (78.203 - 84.757) | 79.170 (76.913 - 81.427) | 80.010 (77.055 - 82.965) |
| GPT-o3-mini | Micro Recall | 86.973 (84.621 - 89.325) | 85.202 (82.839 - 87.564) | 82.925 (80.212 - 85.638) | 83.620 (80.898 - 86.343) | 87.416 (85.312 - 89.519) | 85.139 (82.762 - 87.515) | 82.672 (79.967 - 85.377) | 83.873 (81.474 - 86.273) | 87.045 (84.330 - 89.760) | 85.575 (82.948 - 88.202) | 80.640 (77.413 - 83.867) | 82.215 (79.202 - 85.228) | 86.625 (84.057 - 89.193) | 84.735 (81.843 - 87.627) | 80.010 (76.823 - 83.197) | 81.375 (77.869 - 84.881) |
| GPT-4o-mini | Cost | 0.090 (0.077 - 0.104) | 0.496 (0.355 - 0.637) | 0.033 (0.032 - 0.034) | 0.481 (0.340 - 0.622) | 0.092 (0.078 - 0.105) | 0.497 (0.356 - 0.638) | 0.034 (0.034 - 0.035) | 0.482 (0.341 - 0.623) | 0.054 (0.046 - 0.063) | 0.483 (0.342 - 0.624) | 0.020 (0.020 - 0.020) | 0.474 (0.333 - 0.615) | 0.055 (0.047 - 0.064) | 0.484 (0.343 - 0.625) | 0.021 (0.021 - 0.021) | 0.475 (0.334 - 0.616) |
| GPT-o3-mini | Cost | 0.663 (0.563 - 0.763) | 0.703 (0.562 - 0.845) | 0.242 (0.237 - 0.247) | 0.591 (0.450 - 0.733) | 0.673 (0.573 - 0.773) | 0.713 (0.572 - 0.855) | 0.252 (0.247 - 0.257) | 0.601 (0.460 - 0.743) | 0.400 (0.339 - 0.460) | 0.608 (0.467 - 0.749) | 0.147 (0.143 - 0.150) | 0.541 (0.400 - 0.682) | 0.407 (0.346 - 0.467) | 0.615 (0.474 - 0.756) | 0.154 (0.151 - 0.157) | 0.548 (0.407 - 0.689) |
| GPT-4o-mini | Time | 277.467 (-15.504 - 570.438) | 698.952 (525.928 - 871.977) | 127.446 (118.783 - 136.109) | 689.129 (517.946 - 860.311) | 134.343 (124.561 - 144.125) | 697.827 (528.618 - 867.037) | 180.095 (82.832 - 277.358) | 686.546 (514.642 - 858.451) | 85.571 (78.989 - 92.152) | 649.812 (476.901 - 822.723) | 79.595 (73.649 - 85.542) | 643.515 (471.790 - 815.240) | 84.092 (77.564 - 90.621) | 644.416 (472.263 - 816.569) | 73.316 (67.722 - 78.911) | 641.975 (470.013 - 813.938) |
| GPT-o3-mini | Time | 649.576 (579.184 - 719.968) | 1142.825 (968.078 - 1317.572) | 582.214 (531.811 - 632.616) | 1205.428 (1006.966 - 1403.889) | 648.587 (587.662 - 709.512) | 1238.629 (1050.032 - 1427.227) | 1010.154 (419.007 - 1601.301) | 1173.793 (990.654 - 1356.932) | 384.252 (339.330 - 429.175) | 913.216 (742.118 - 1084.314) | 359.350 (323.674 - 395.026) | 947.115 (765.343 - 1128.888) | 390.282 (347.155 - 433.409) | 947.736 (778.357 - 1117.114) | 538.781 (235.208 - 842.354) | 944.620 (764.310 - 1124.930) |

**Supplementary Table 4**. Classification performance and operative metrics of the zero-shot, no RAG retrieval prompt method and few-shot, no RAG retrieval prompt method applied to the reasoning (o1, o3-mini) and generalist (gpt-4o, gpt-4o-mini) LLM on the CONSORT Test dataset split from the SPIRIT-CONSORT-TM corpus. Metrics are shown for all 83 CONSORT and SPIRIT checklist items and the 79 CONSORT-specific checklist items reported in the SPIRIT-CONSORT-TM corpus.

| Model | Metric | Zero-shot All Item | Few-shot All Item | Zero-shot CONSORT Item | Few-shot CONSORT Item |
|---|---|---|---|---|---|
| GPT-4o | Accuracy | 82.925 (80.034 - 85.816) | 85.771 (83.593 - 87.949) | 84.630 (81.392 - 87.868) | 85.575 (82.707 - 88.443) |
| GPT-4o-mini | Accuracy | 78.307 (76.378 - 80.237) | 81.976 (79.849 - 84.103) | 80.535 (77.678 - 83.392) | 82.950 (79.785 - 86.115) |
| GPT-o1 | Accuracy | 89.006 (86.371 - 91.641) | 90.271 (87.968 - 92.574) | 91.650 (88.369 - 94.931) | 91.650 (88.734 - 94.566) |
| GPT-o3-mini | Accuracy | 89.123 (87.123 - 91.124) | 89.693 (87.634 - 91.752) | 90.090 (87.204 - 92.976) | 89.775 (86.837 - 92.713) |
| GPT-4o | Macro F1 | 81.492 (78.382 - 84.602) | 84.716 (82.379 - 87.053) | 79.237 (75.533 - 82.940) | 80.912 (77.500 - 84.324) |
| GPT-4o-mini | Macro F1 | 76.365 (74.417 - 78.312) | 80.672 (78.567 - 82.776) | 74.430 (71.627 - 77.233) | 77.877 (74.893 - 80.862) |
| GPT-o1 | Macro F1 | 88.108 (85.118 - 91.098) | 89.592 (87.170 - 92.013) | 89.305 (85.331 - 93.280) | 89.485 (86.014 - 92.957) |
| GPT-o3-mini | Macro F1 | 88.369 (86.435 - 90.303) | 88.944 (86.797 - 91.090) | 87.342 (84.434 - 90.251) | 87.054 (83.951 - 90.157) |
| GPT-4o | Micro F1 | 82.925 (80.034 - 85.816) | 85.771 (83.593 - 87.949) | 84.630 (81.392 - 87.868) | 85.575 (82.707 - 88.443) |
| GPT-4o-mini | Micro F1 | 78.307 (76.378 - 80.237) | 81.976 (79.849 - 84.103) | 80.535 (77.678 - 83.392) | 82.950 (79.785 - 86.115) |
| GPT-o1 | Micro F1 | 89.006 (86.371 - 91.641) | 90.271 (87.968 - 92.574) | 91.650 (88.369 - 94.931) | 91.650 (88.734 - 94.566) |
| GPT-o3-mini | Micro F1 | 89.123 (87.123 - 91.124) | 89.693 (87.634 - 91.752) | 90.090 (87.204 - 92.976) | 89.775 (86.837 - 92.713) |
| GPT-4o | Macro Precision | 85.603 (83.390 - 87.816) | 86.720 (84.833 - 88.606) | 87.625 (84.491 - 90.759) | 86.034 (83.008 - 89.060) |
| GPT-4o-mini | Macro Precision | 81.449 (80.262 - 82.635) | 83.668 (81.878 - 85.457) | 81.615 (78.854 - 84.375) | 85.169 (82.080 - 88.259) |

| | | | | | |
|---|---|---|---|---|---|
| GPT-o1 | Macro Precision | 89.262 (86.872 - 91.652) | 90.050 (87.932 - 92.168) | 92.742 (89.419 - 96.066) | 91.771 (88.828 - 94.715) |
| GPT-o3-mini | Macro Precision | 89.992 (88.137 - 91.848) | 89.794 (87.867 - 91.722) | 91.211 (88.633 - 93.790) | 89.601 (86.652 - 92.550) |
| GPT-4o | Micro Precision | 82.925 (80.034 - 85.816) | 85.771 (83.593 - 87.949) | 84.630 (81.392 - 87.868) | 85.575 (82.707 - 88.443) |
| GPT-4o-mini | Micro Precision | 78.307 (76.378 - 80.237) | 81.976 (79.849 - 84.103) | 80.535 (77.678 - 83.392) | 82.950 (79.785 - 86.115) |
| GPT-o1 | Micro Precision | 89.006 (86.371 - 91.641) | 90.271 (87.968 - 92.574) | 91.650 (88.369 - 94.931) | 91.650 (88.734 - 94.566) |
| GPT-o3-mini | Micro Precision | 89.123 (87.123 - 91.124) | 89.693 (87.634 - 91.752) | 90.090 (87.204 - 92.976) | 89.775 (86.837 - 92.713) |
| GPT-4o | Macro Recall | 82.823 (79.625 - 86.021) | 85.497 (82.838 - 88.157) | 78.265 (74.411 - 82.118) | 80.008 (76.301 - 83.716) |
| GPT-4o-mini | Macro Recall | 78.083 (75.786 - 80.379) | 81.797 (79.403 - 84.192) | 74.223 (71.409 - 77.038) | 77.118 (73.945 - 80.290) |
| GPT-o1 | Macro Recall | 88.604 (85.340 - 91.869) | 89.947 (87.286 - 92.609) | 88.119 (83.887 - 92.351) | 88.623 (84.945 - 92.301) |
| GPT-o3-mini | Macro Recall | 88.942 (86.895 - 90.988) | 89.338 (87.063 - 91.613) | 86.156 (83.156 - 89.156) | 86.153 (82.986 - 89.321) |
| GPT-4o | Micro Recall | 82.925 (80.034 - 85.816) | 85.771 (83.593 - 87.949) | 84.630 (81.392 - 87.868) | 85.575 (82.707 - 88.443) |
| GPT-4o-mini | Micro Recall | 78.307 (76.378 - 80.237) | 81.976 (79.849 - 84.103) | 80.535 (77.678 - 83.392) | 82.950 (79.785 - 86.115) |
| GPT-o1 | Micro Recall | 89.006 (86.371 - 91.641) | 90.271 (87.968 - 92.574) | 91.650 (88.369 - 94.931) | 91.650 (88.734 - 94.566) |
| GPT-o3-mini | Micro Recall | 89.123 (87.123 - 91.124) | 89.693 (87.634 - 91.752) | 90.090 (87.204 - 92.976) | 89.775 (86.837 - 92.713) |
| GPT-4o | Cost | 1.853 (1.623 - 2.083) | 1.876 (1.646 - 2.106) | 1.217 (1.057 - 1.376) | 1.233 (1.073 - 1.393) |
| GPT-4o-mini | Cost | 0.346 (0.283 - 0.410) | 0.348 (0.284 - 0.412) | 0.308 (0.249 - 0.368) | 0.309 (0.250 - 0.369) |
| GPT-o1 | Cost | 9.337 (7.800 - 10.874) | 9.475 (7.938 - 11.012) | 5.721 (4.766 - 6.676) | 5.819 (4.863 - 6.774) |
| GPT-o3-mini | Cost | 0.956 (0.825 - 1.086) | 0.966 (0.835 - 1.097) | 0.676 (0.576 - 0.775) | 0.683 (0.583 - 0.783) |

| | | | | | |
|---|---|---|---|---|---|
| GPT-4o | Time | 493.148 (402.463 - 583.832) | 479.662 (393.786 - 565.538) | 405.529 (318.383 - 492.675) | 401.810 (318.408 - 485.212) |
| GPT-4o-mini | Time | 400.404 (317.192 - 483.616) | 402.058 (318.624 - 485.491) | 349.822 (267.194 - 432.450) | 352.307 (269.739 - 434.875) |
| GPT-o1 | Time | 1295.000 (1135.345 - 1454.654) | 1321.871 (1162.034 - 1481.709) | 868.741 (725.497 - 1011.985) | 894.408 (751.034 - 1037.782) |
| GPT-o3-mini | Time | 850.518 (707.386 - 993.650) | 1334.997 (354.151 - 2315.844) | 617.259 (499.517 - 735.000) | 619.226 (502.551 - 735.901) |

**Supplementary Table 5**. Classification performance and operative metrics of the zero-shot, no RAG retrieval prompt method and few-shot, no RAG retrieval prompt method applied to the reasoning (o1, o3-mini) and generalist (gpt-4o, gpt-4o-mini) LLM on the SPIRIT Test dataset split from the SPIRIT-CONSORT-TM corpus. Metrics are shown for all 83 CONSORT and SPIRIT checklist items and the 72 SPIRIT-specific checklist items reported in the SPIRIT-CONSORT-TM corpus.

| Model | Metric | Zero-shot All Item | Few-shot All Item | Zero-shot SPIRIT Item | Few-shot SPIRIT Item |
|---|---|---|---|---|---|
| GPT-4o | Accuracy | 83.557 (81.856 - 85.259) | 86.846 (85.367 - 88.325) | 86.978 (85.023 - 88.933) | 89.485 (87.945 - 91.025) |
| GPT-4o-mini | Accuracy | 81.533 (79.523 - 83.543) | 82.608 (80.667 - 84.549) | 83.451 (81.325 - 85.578) | 84.549 (82.248 - 86.849) |
| GPT-o1 | Accuracy | 88.825 (86.756 - 90.895) | 92.169 (89.897 - 94.440) | 90.336 (87.913 - 92.759) | 92.575 (90.053 - 95.096) |
| GPT-o3-mini | Accuracy | 90.136 (88.314 - 91.957) | 91.654 (90.316 - 92.991) | 92.071 (89.892 - 94.250) | 93.403 (91.817 - 94.989) |
| GPT-4o | Macro F1 | 82.652 (81.002 - 84.303) | 86.180 (84.658 - 87.702) | 85.124 (83.263 - 86.985) | 88.243 (86.769 - 89.718) |
| GPT-4o-mini | Macro F1 | 80.481 (78.224 - 82.737) | 81.759 (79.643 - 83.875) | 81.117 (78.690 - 83.544) | 82.711 (80.157 - 85.264) |
| GPT-o1 | Macro F1 | 88.285 (86.032 - 90.538) | 91.810 (89.439 - 94.182) | 89.155 (86.648 - 91.662) | 91.750 (89.230 - 94.271) |
| GPT-o3-mini | Macro F1 | 89.638 (87.790 - 91.486) | 91.206 (89.816 - 92.597) | 91.077 (89.033 - 93.121) | 92.481 (90.850 - 94.113) |
| GPT-4o | Micro F1 | 83.557 (81.856 - 85.259) | 86.846 (85.367 - 88.325) | 86.978 (85.023 - 88.933) | 89.485 (87.945 - 91.025) |
| GPT-4o-mini | Micro F1 | 81.533 (79.523 - 83.543) | 82.608 (80.667 - 84.549) | 83.451 (81.325 - 85.578) | 84.549 (82.248 - 86.849) |
| GPT-o1 | Micro F1 | 88.825 (86.756 - 90.895) | 92.169 (89.897 - 94.440) | 90.336 (87.913 - 92.759) | 92.575 (90.053 - 95.096) |
| GPT-o3-mini | Micro F1 | 90.136 (88.314 - 91.957) | 91.654 (90.316 - 92.991) | 92.071 (89.892 - 94.250) | 93.403 (91.817 - 94.989) |
| GPT-4o | Macro Precision | 86.164 (84.416 - 87.912) | 87.657 (85.905 - 89.408) | 88.597 (86.769 - 90.426) | 89.694 (88.005 - 91.383) |
| GPT-4o-mini | Macro Precision | 83.587 (81.851 - 85.324) | 83.717 (81.944 - 85.489) | 84.296 (82.177 - 86.415) | 84.602 (81.894 - 87.310) |

| | | | | | |
|---|---|---|---|---|---|
| GPT-o1 | Macro Precision | 89.316 (87.130 - 91.501) | 92.031 (89.629 - 94.434) | 90.437 (88.102 - 92.773) | 92.272 (89.905 - 94.638) |
| GPT-o3-mini | Macro Precision | 90.984 (88.887 - 93.080) | 91.780 (90.200 - 93.359) | 92.853 (90.681 - 95.024) | 93.355 (91.742 - 94.968) |
| GPT-4o | Micro Precision | 83.557 (81.856 - 85.259) | 86.846 (85.367 - 88.325) | 86.978 (85.023 - 88.933) | 89.485 (87.945 - 91.025) |
| GPT-4o-mini | Micro Precision | 81.533 (79.523 - 83.543) | 82.608 (80.667 - 84.549) | 83.451 (81.325 - 85.578) | 84.549 (82.248 - 86.849) |
| GPT-o1 | Micro Precision | 88.825 (86.756 - 90.895) | 92.169 (89.897 - 94.440) | 90.336 (87.913 - 92.759) | 92.575 (90.053 - 95.096) |
| GPT-o3-mini | Micro Precision | 90.136 (88.314 - 91.957) | 91.654 (90.316 - 92.991) | 92.071 (89.892 - 94.250) | 93.403 (91.817 - 94.989) |
| GPT-4o | Macro Recall | 84.079 (82.678 - 85.480) | 87.037 (85.659 - 88.415) | 84.930 (82.998 - 86.862) | 88.125 (86.551 - 89.699) |
| GPT-4o-mini | Macro Recall | 82.051 (79.696 - 84.405) | 83.068 (80.723 - 85.414) | 81.343 (78.673 - 84.012) | 82.980 (80.268 - 85.692) |
| GPT-o1 | Macro Recall | 89.225 (87.239 - 91.211) | 92.561 (90.803 - 94.320) | 89.083 (86.663 - 91.502) | 91.860 (89.556 - 94.163) |
| GPT-o3-mini | Macro Recall | 90.489 (89.079 - 91.898) | 91.908 (90.672 - 93.145) | 90.839 (88.955 - 92.723) | 92.474 (90.645 - 94.303) |
| GPT-4o | Micro Recall | 83.557 (81.856 - 85.259) | 86.846 (85.367 - 88.325) | 86.978 (85.023 - 88.933) | 89.485 (87.945 - 91.025) |
| GPT-4o-mini | Micro Recall | 81.533 (79.523 - 83.543) | 82.608 (80.667 - 84.549) | 83.451 (81.325 - 85.578) | 84.549 (82.248 - 86.849) |
| GPT-o1 | Micro Recall | 88.825 (86.756 - 90.895) | 92.169 (89.897 - 94.440) | 90.336 (87.913 - 92.759) | 92.575 (90.053 - 95.096) |
| GPT-o3-mini | Micro Recall | 90.136 (88.314 - 91.957) | 91.654 (90.316 - 92.991) | 92.071 (89.892 - 94.250) | 93.403 (91.817 - 94.989) |
| GPT-4o | Cost | 1.765 (1.404 - 2.126) | 1.788 (1.427 - 2.149) | 1.473 (1.162 - 1.784) | 1.493 (1.182 - 1.804) |
| GPT-4o-mini | Cost | 0.342 (0.225 - 0.459) | 0.344 (0.227 - 0.461) | 0.325 (0.211 - 0.439) | 0.326 (0.212 - 0.440) |
| GPT-o1 | Cost | 9.299 (6.863 - 11.735) | 9.437 (7.001 - 11.873) | 7.557 (5.562 - 9.552) | 7.674 (5.679 - 9.668) |
| GPT-o3-mini | Cost | 0.917 (0.702 - 1.133) | 0.927 (0.712 - 1.143) | 0.789 (0.596 - 0.982) | 0.798 (0.604 - 0.991) |

| | | | | | |
|---|---|---|---|---|---|
| GPT-4o | Time | 469.928 (411.147 - 528.709) | 470.816 (412.189 - 529.442) | 431.443 (374.238 - 488.648) | 434.252 (376.444 - 492.060) |
| GPT-4o-mini | Time | 385.522 (334.167 - 436.877) | 383.259 (329.947 - 436.571) | 362.319 (310.827 - 413.812) | 360.518 (307.705 - 413.330) |
| GPT-o1 | Time | 1318.066 (1202.311 - 1433.821) | 1342.920 (1211.672 - 1474.168) | 1085.041 (983.167 - 1186.915) | 1106.616 (992.211 - 1221.021) |
| GPT-o3-mini | Time | 734.534 (666.279 - 802.789) | 756.343 (680.876 - 831.809) | 633.377 (569.126 - 697.628) | 650.981 (583.698 - 718.264) |

**Supplementary Table 6**. Examples of failure modes observed by AutoReporter.

| Failure Mode | PRISMA/CONSORT/SPIRIT Checklist Item | Reference Rating | Incorrect AutoReporter Rating | Explanation of Failure Mode |
|---|---|---|---|---|
| **Missed Table/Figure Content** | PRISMA Item 17 (Study selection – provide numbers of studies screened and included, ideally with a flow diagram) | **Fully Reported** – "Figure 1 presents the PRISMA flow diagram showing records identified (n=1,204), screened (n=1,000), excluded (n=850), and included (n=150)." | **Not Fully Reported** | AutoReporter overlooked the flow diagram in the figure and flagged the item as "Not Fully Reported" because it only parsed narrative text. |
| **Composite Item Ambiguity** | CONSORT Item 4a (Eligibility criteria for participants and settings where data were collected) | **Fully Reported** – "Participants were adults aged 18–65 with type 2 diabetes, recruited from three urban clinics in Toronto." | **Not Fully Reported** | The model correctly identified eligibility criteria but failed to integrate setting details, producing an inconsistent "Not Fully Reported" rating for the composite item. |
| **Ambiguously Phrased Item** | SPIRIT Item 23 (Plans for dissemination, including access to full protocol, participant-level dataset, and statistical code) | **Not Fully Reported** – "Results will be published in peer-reviewed journals." (No mention of dataset or code availability.) | **Fully Reported** | Because the item contains multiple sub-components, AutoReporter over-generalized from partial reporting and incorrectly marked the item as "Fully Reported". |
| **Misattributed Quoted Evidence** | PRISMA Item 8 (Full electronic search strategy for at least one database) | **Not Fully Reported** – "Search terms included 'cancer' AND 'radiotherapy' …" (lacked full reproducible search strings). | **Fully Reported** – AutoReporter quoted a nearby sentence: "We searched PubMed and Embase for all eligible studies …" | Semantically similar sentences in the same section led AutoReporter to select an adjacent but incomplete line as evidence, mis-attributing reporting sufficiency. |
